# Effects of non-pharmaceutical interventions on COVID-19: A Tale of Three Models

**DOI:** 10.1101/2020.07.22.20160341

**Authors:** Vincent Chin, John P.A. Ioannidis, Martin A. Tanner, Sally Cripps

## Abstract

**Objective:** To compare the inference regarding the effectiveness of the various non-pharmaceutical interventions (NPIs) for COVID-19 obtained from different SIR models.

**Study design and setting:** We explored two models developed by Imperial College that considered only NPIs without accounting for mobility (model 1) or only mobility (model 2), and a model accounting for the combination of mobility and NPIs (model 3). Imperial College applied models 1 and 2 to 11 European countries and to the USA, respectively. We applied these models to 14 European countries (original 11 plus another 3), over two different time horizons.

**Results:** While model 1 found that lockdown was the most effective measure in the original 11 countries, model 2 showed that lockdown had little or no benefit as it was typically introduced at a point when the time-varying reproductive number was already very low. Model 3 found that the simple banning of public events was beneficial, while lockdown had no consistent impact. Based on Bayesian metrics, model 2 was better supported by the data than either model 1 or model 3 for both time horizons.

**Conclusions:** Inferences on effects of NPIs are non-robust and highly sensitive to model specification. Claimed benefits of lockdown appear grossly exaggerated.

## 1 Introduction

Until effective and safe vaccines can become widely available, the levers of policy makers to manage COVID-19 have included non-pharmaceutical interventions (NPI), such as social distancing mandates, travel restrictions, self-isolation, banning of public events, closure of schools, and ultimately complete lockdown. These measures aim to reduce infections by decreasing contact between individuals. Given that multiple NPIs are often introduced in quick succession, it is difficult to separate their effects.

Here, we compare the inferences regarding the effectiveness of various NPIs obtained from different SIR (susceptible-infected-removed) models. The first model (model 1) was produced by the Imperial College COVID-19 Response Team and led to arguably the most influential publication to-date in support of large benefits from total lockdown^1^. Its publication in *Nature* ^1^, concluded that complete lockdown was *responsible for 80% of the reduction in the time-varying reproduction number, R*_*t*_, and that 3.1 million deaths were avoided in 11 European countries due to lockdown. The Imperial College team also developed and applied a different model (model 2) to the USA^2^, which assumes *R*_*t*_ varies as a function of mobility. In model 2, there is no explicit causal link between an NPI and *R*_*t*_–NPIs enter the model indirectly via their effects on mobility. Inference regarding the (complex) impact of NPIs is possible by observing the *R*_*t*_ trajectory at the time of intervention/s.

We compare the results and performance (fit to the data) of these two models, when applied to the original 11 countries, plus another 3 European countries for which data were available but had not been included in the original publication. We also considered a third model (model 3), a hydrid of the first two, that considers both mobility and various NPIs together. We aim to understand if inferences are robust to model specification and whether some model provides a better fit than others.

## 2 Methods

### Data

We compare the impact of NPIs and mobility on *R*_*t*_ for three models, two time horizons and two sets of European countries. Specifically,

1. For all models, we examine the evolution of *R*_*t*_ for two time horizons: up to May 5th (the end date chosen by Flaxman et al. ^1^), and July 12th to allow investigating both the imposition and lifting of various NPIs.
2. The original publication by Flaxman et al.^1^ had included 11 European countries (Austria, Belgium, Denmark, France, Germany, Italy, Norway, Spain, Sweden, Switzerland, United Kingdom). However, suitable data were also available for the Netherlands, Portugal, and Greece; therefore we also considered 14 countries.

Seeding of new infections in all models was chosen to be 10 days before the day a given country has cumulatively observed 10 deaths so that mobility data are available for all countries examined and thus allow a fair comparison between models. Flaxman et al. ^1^ chose the seeding of new infections to be 30 days before a country has cumulatively observed 10 deaths. We have altered the prior for the initial infection count, to reflect this modification. Seeding dates appear in Table A.1.

For mobility data we follow Unwin et al.^2^, and use Google’s COVID-19 Community Mobility Report^3^, which provides data measuring the percentage change in mobility compared to a baseline level for visits to: retailers and recreation venues, grocery markets and pharmacies, parks, transit stations, workplaces and residential places. We use the average change in mobility across all locations, excluding residential places and parks. Mobility indicators are proxies for changes in human behavior and of exposure risk. Behavior change could be due to one or more centrally imposed interventions or the product of individuals responding to the epidemic on their own initiative.

### Model 1 (all NPIs considered)

In model 1, the evolution of *R*_*t*_ is given by,

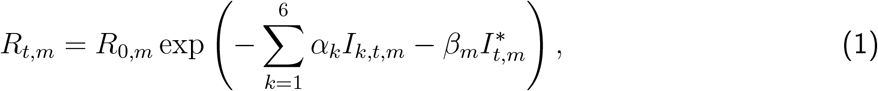

where *R*_*t,m*_ is the effective reproductive rate for country *m* at time *t* and *I*_*k,t,m*_ is an indicator variable, where *I*_*k,t,m*_ = 1 if NPI *k* is in place at time *t*, for country *m* and *I*_*k,t,m*_ = 0 otherwise, for *k* = 1, …, 6. The subscript *k* refers to the various NPIs (Table A.2). The variable 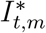 indicates the last intervention. In all countries except Sweden, this was lockdown, see Flaxman et al. ^1^ for details. We allow the impact of lifting an NPI on *R*_*t*_ to be different in magnitude from the impact of imposing that NPI in the first place. The timing of lifting NPIs in different countries appears in Table A.3.

In Equation 1, the proportional variation of *R*_*t*_ from the initial *R*_0_ is modelled as a step function and only allowed to change, immediately so, in response to an intervention. Therefore, any decrease in *R*_*t*_ (even if this decrease is a result of the increasing proportion of the population who are infected, changes in human behavior, clustered contact structures and/or pre-existing immunity^4^) must, by construction, be attributed to interventions; the impact of a new intervention is immediate without time lag or gradual change. This assumption was clearly made for simplicity but is unrealistic.

### Model 2 (Mobility Only Considered)

In model 2, the proportional variation of *R*_*t*_ from *R*_0_ is allowed to vary with mobility. Model 2 does not presume *R*_*t*_ follows a step function and is therefore capable of capturing more gradual changes over time. The impact of mobility on *R*_*t*_ is allowed to vary across countries by use of country-specific random effects terms. Specifically,

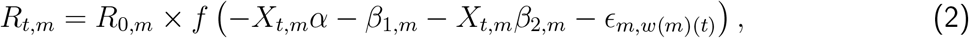

where 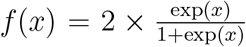 is twice the inverse of the logit function, *X*_*t,m*_ is the average change in mobility, excluding residential places and parks, at time *t* for country *m* and *E*_*m,w*(*m*)(*t*)_ is a weekly AR(2) process centered around zero. In Equation 2, *α* is a measure of the impact of the average change in mobility on *R*_*t*_ which is common to all countries, while *β*_2,*m*_ measures country-specific deviations from this common value. The advantage of model 2 is that it gives a more flexible estimate of *R*_*t*_, allowing it to change with mobility trends. Although NPIs are not explicitly included in the model, the impact of an NPI can be measured by observing the value of *R*_*t*_, and its subsequent change, when specific interventions were imposed.

### Model 3 (Mobility and NPIs jointly considered)

After communication with the Imperial College team, we also consider a third model (model 3) which jointly includes mobility and NPIs. The motivation behind the formulation of model 3 is to attempt to untangle the impacts of mobility, lockdown and other NPIs. In our communication, the Imperial College team proposed a similar model but only included mobility and a single NPI, lockdown, in their model. Given that our goal is to quantify the relative contributions of several NPIs, we consider all NPIs, plus mobility.

However, we caution against using model 3 as a tool for inference. NPIs may impact mobility in possibly non-linear, non-additive, lagged and interactive fashions, with possibly complex feedback. We have included this model here to compare its performance against models 1 and 2. The functional form of *R*_*t,m*_, in model 3 is:

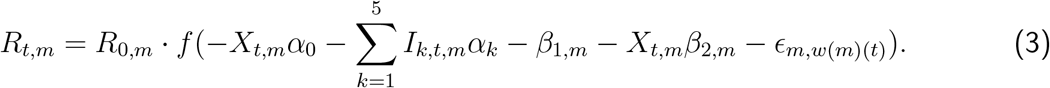

For a discussion of prior specification and Bayesian measures of model fit for all models, see Appendix B.

## 3 Results

### Mobility

Figure 1 and Figure A.1 show that for most countries the initial reduction in mobility preceded the date of the first lockdown. This suggests that people’s behavior changed in response to earlier, less severe interventions such as banning of public events and social distancing, and/or as a result of individual choices in the face of an unknown, but potentially catastrophic, pandemic.

**Figure 1:**
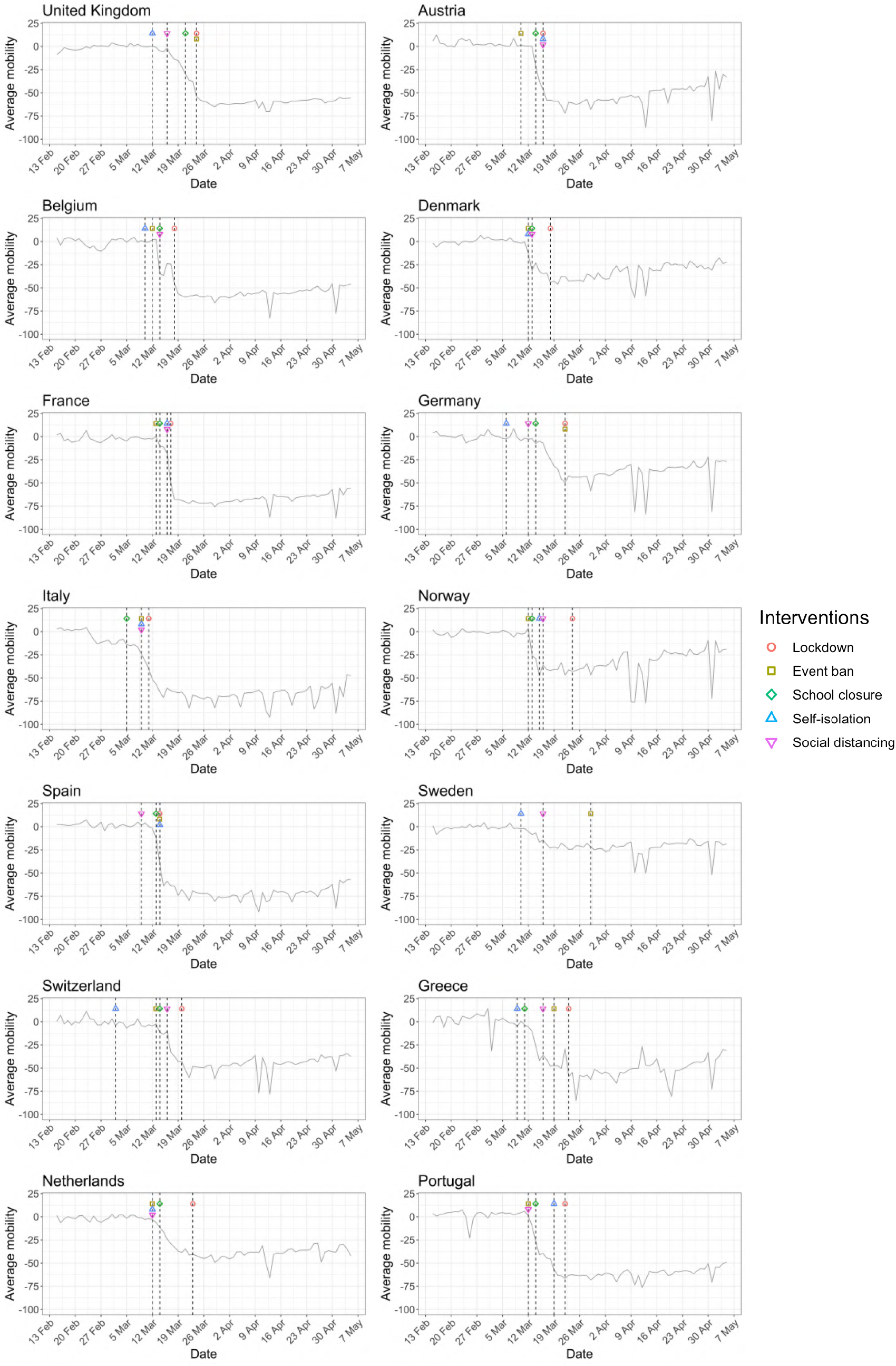
Percentage change in average mobility from baseline level from February 15th to May 5th in each of the European countries examined in Flaxman et al. ^1^, as well as an additional three countries consisting of Greece, the Netherlands and Portugal.

### Convergence diagnostics

Convergence diagnostics (trace plots and 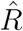^5^ based on 10 chains - see also^6^) for all three models and both time horizons appear in Figure A.2, providing strong evidence that the Markov chains have converged.

### Comparison of models up to May 5th

While model 1 and model 2 give very different trajectories of *R*_*t*_ (Figures 2a, A.3a–A.12a, Appendix C), both models produce visually similar fits to the observed daily death counts, i.e. different trajectories of *R*_*t*_ may give rise to the same data and hence different inference surrounding the impact of various NPIs.

**Figure 2:**
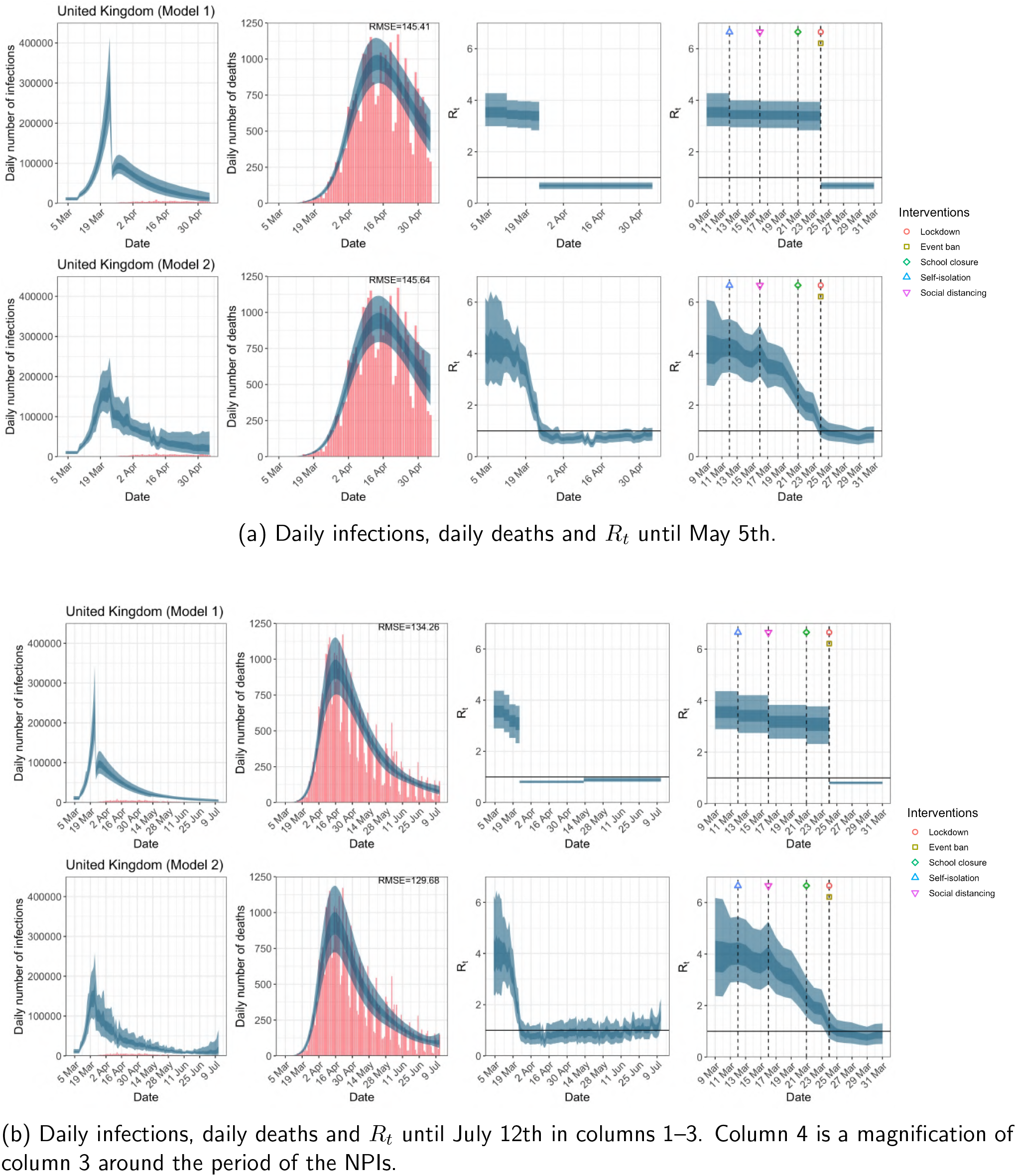
United Kingdom. The start time for the plots is 10 days before 10 deaths are recorded. Observed counts of daily infections and daily deaths are shown in red, and their corresponding 50% and 95% CIs are shown in dark blue and light blue respectively.

For the 11 countries (Table 1), the inference from model 1 indicates that lockdown had the biggest impact of all the interventions in all countries with an average reduction in *R*_*t*_ of 80%. In contrast, model 2 shows clearly that *R*_*t*_ was falling well before lockdown, excluding Sweden that had no lockdown. In the other 10 countries, *R*_*t*_ *<* 1.0 at the time of lockdown in 4 countries and only 1.0 − 1.3 in another 3 countries (all three 95% CIs contained 1.0).

**Table 1:** Comparison of the value of *R*_*t*_ at lockdown (LD) and its 95% CIs between model 1 and 2 for all eleven countries analysed in Flaxman et al. ^1^ for the time horizon March 4th to May 5th. Values of basic reproduction number *R*_0_ and *R*_*t*_ immediately after the introduction of other NPIs for both models are given in Table A.5 in the Appendix.

| Country | Model 1 |  |  | Model 2 |
| --- | --- | --- | --- | --- |
| | $R_t$ one day before LD | $R_t$ at LD | % change | $R_t$ at LD |
| UK | 3.39<br>(2.84,3.94) | 0.68<br>(0.55,0.81) | -79.67<br>(-85.29, -72.96) | 1.11<br>(0.75,1.60) |
| Austria | 2.96<br>(1.67,4.50) | 0.52<br>(0.40,0.64) | -81.42<br>(-88.80, -69.47) | 0.87<br>(0.42,1.55) |
| Belgium | 4.30<br>(2.87,6.06) | 0.90<br>(0.78,1.02) | -78.31<br>(-85.99, -67.26) | 4.83<br>(3.47,6.45) |
| Denmark | 3.25<br>(1.98,4.81) | 0.68<br>(0.57,0.80) | -78.11<br>(-86.01, -65.70) | 0.58<br>(0.28,1.05) |
| France | 4.06<br>(2.98,4.95) | 0.71<br>(0.61,0.82) | -82.08<br>(-87.07, -74.21) | 1.69<br>(1.16,2.39) |
| Germany | 3.68<br>(2.94,4.51) | 0.73<br>(0.60,0.85) | -79.99<br>(-85.84, -72.48) | 1.02<br>(0.68,1.47) |
| Italy | 2.90<br>(2.17,3.46) | 0.70<br>(0.63,0.78) | -75.35<br>(-80.98, -66.51) | 1.30<br>(0.86,1.76) |
| Norway | 2.42<br>(1.36,3.71) | 0.40<br>(0.25,0.57) | -82.30<br>(-91.04, -69.16) | 0.50<br>(0.27,0.79) |
| Spain | 4.29<br>(3.35,5.39) | 0.67<br>(0.59,0.75) | -84.05<br>(-88.43, -78.72) | 1.78<br>(1.22,2.42) |
| Sweden | — | — | — | — |
| Switzerland | 2.67<br>(1.93,3.48) | 0.55<br>(0.44,0.68) | -78.61<br>(-86.43, -67.32) | 0.93<br>(0.62,1.31) |

When we considered 3 additional countries (Table C.1), the average reduction in *R*_*t*_ from lockdown shrank to 73% in model 1. Model 2 shows *R*_*t*_ *<* 1.0 in 7 countries and 1.0−1.3 in another 3 countries when lockdown was imposed. In particular, the three added countries already had *R*_*t*_ *<* 1.0 at the time of lockdown. For Greece and Portugal, *R*_*t*_ was already so low (0.34 and 0.67, respectively) that even the 95% CIs excluded 1.0.

Model 3 provides different inference yet again. Only the mobility and banning of public events have 95% CIs for regression coefficients which do not include zero (Figure A.2). The impact of lockdown is not statistically significant (95% CI is − 0.23, 4.25).

In comparing the models, Table A.4 shows that model 2 provides a lower RMSE for eight of the eleven original countries considered by Flaxman et al. ^1^, for the period March 4th to May 5th. The three countries for which model 1 had a lower RMSE were the UK, Germany and Norway.

Table 2 demonstrates that model 2 is best supported by the data for all three information criteria, WAIC1, WAIC2 and DIC (see Appendix B.2). Model 3 is the next best supported by the data, while model 1 published in Nature is the least supported.

**Table 2:** Estimates and standard errors of various information criteria; the Watanabe-Akaike information criterion, *WAIC*1 = −2lppd + 2*p*_*WAIC*1_ and *WAIC*2 = −2lppd + 2*p*_*WAIC*2_ which uses lppd as a measure of fit with *p*_*W AIC*1_ and *p*_*WAIC*2_ as the effective number of parameters to penalize the fit respectively; the Deviance information criterion 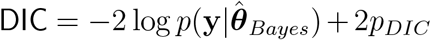 which uses 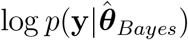, as the measure of fit, and *p*_*DIC*_ as the penalty. Note that a lower value implies a better predictive density, and the preferred model for each criteria and time period is shown in bold. See Appendix B for computational details.

| Model | Time period | WAIC1 | WAIC2 | DIC |
| --- | --- | --- | --- | --- |
| 1 | Up to May 5th | 5658.31±0.17 | 5662.08±0.19 | 5657.15±0.16 |
| **2** | Up to May 5th | **5626.93±0.18** | **5632.06±0.21** | **5626.71±0.17** |
| 3 | Up to May 5th | 5634.10±0.18 | 5639.48±0.22 | 5633.86±0.17 |
| 1 | Up to July 12th | 13425.12±0.76 | 13483.00±2.54 | 13398.01±0.27 |
| **2** | Up to July 12th | **13370.85±1.59** | **13433.07±2.21** | **13346.05±0.24** |
| 3 | Up to July 12th | 13388.38±1.04 | 13450.76±2.05 | 13363.04±0.24 |

### Comparison of models up to July 12th

The analysis of the time horizon March 4th to July 12th, leads to very similar conclusions (Figures 2b, A.3b–A.12b, A.13–A.15). Table 3 indicates that the impact of lockdown on the relative reduction in *R*_*t*_ was 64% for model 1, while in model 2, 7 countries already had *R*_*t*_ ≤ 1.0 and only two countries had 95% CIs for *R*_*t*_ exceeding 1.0 at the time of lockdown. In model 3, in contrast to the period until May 5th, with longer follow-up lockdown is statistically significant (95% CI is 0.23,1.43)

**Table 3:**
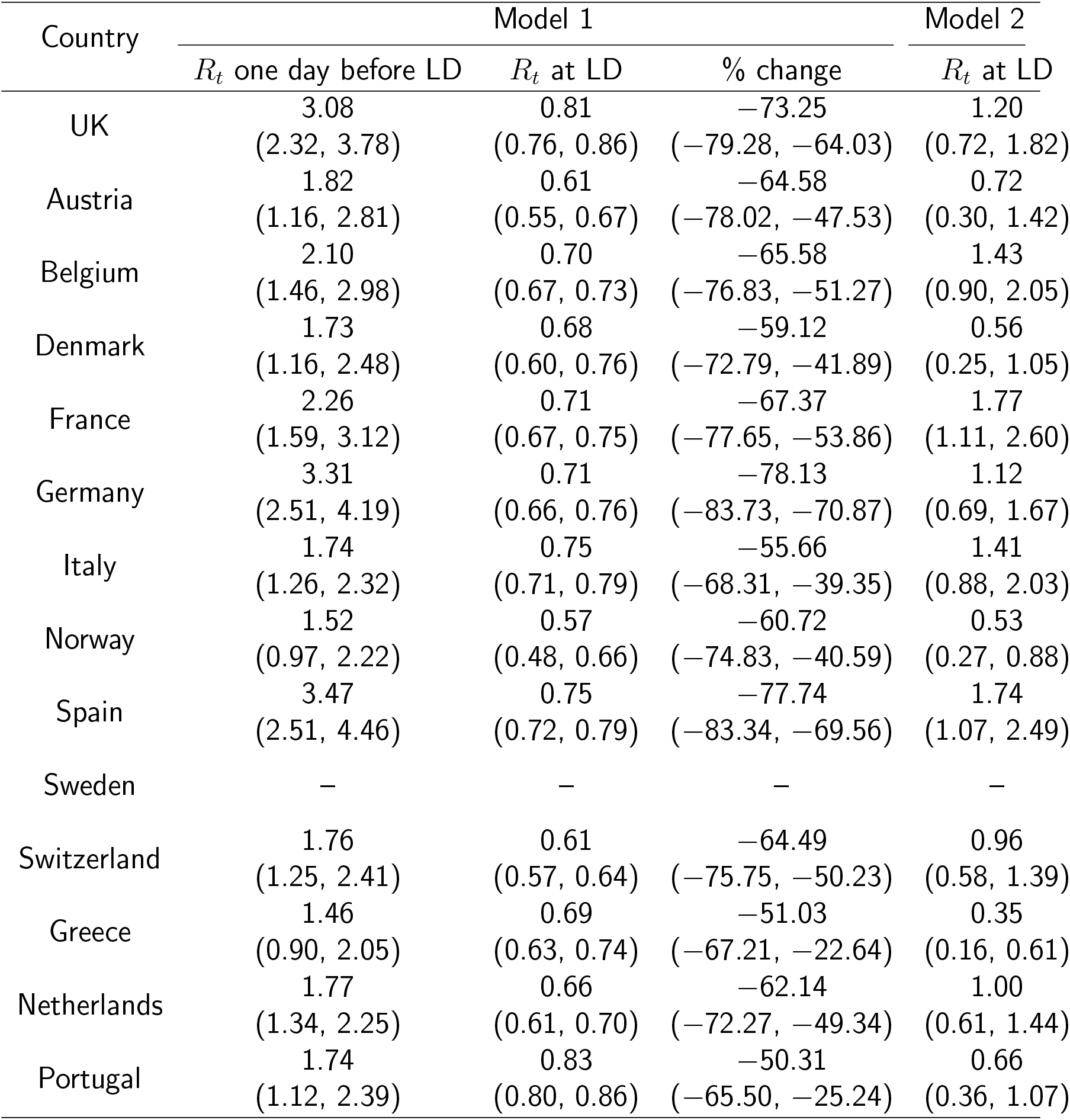
Comparison of the value of *R*_*t*_ at lockdown (LD) and its 95% CIs between model 1 and 2 for all eleven countries analysed in Flaxman et al. ^1^ and an additional three countries of Greece, Netherlands and Portugal, for the time horizon March 4th to July 12th.

In comparing the models, Table A.4 shows that model 2 provides a lower RMSE than model 1 for all countries for the period March 4th to July 12th, except Austria and Norway. Similarly, Table 2 again demonstrates that model 2 is best supported by the data for all three information criteria, WAIC1, WAIC2 and DIC.

### Change of start date

Inferences regarding the impact of the imposition of NPIs is not substantially affected by the start date nor the priors for the initial infection count (Figure A.16).

## 4 Discussion

We demonstrate that effects of NPIs are non-robust and highly sensitive to model specification, assumptions and data employed to fit models. We obtained very different inferences regarding the effectiveness of lockdown measures in terms of curbing the epidemic wave and reducing fatalities. Lockdown appeared the most effective measure to save lives in the original analysis of 11 European countries performed by the Imperial College team through model 1. This analysis was published in *Nature* and has probably had a major impact to maintain a mentality among policy makers that lockdown should be used during the advent of second waves in many countries in the Fall of 2020. However, model 2 (which was also originally developed by the same team), suggests that these impacts were highly exaggerated, with little or no benefit from lockdown in most of the same countries.

Importantly, model 2 typically outperformed model 1 in data fit. Consideration of longer follow-up that included also the lifting of many measures still suggested that the originally^1^ claimed effects of lockdown are grossly overstated. Fitting yet a third model, resulted in yet further variant conclusions, with only mobility and event ban having regression coefficients with 95%CIs that did not contain 0 for the period until May 5th.

The different results and inferences of these models may be partly explained by the highly correlated structure of NPIs and mobility data, as well as the dense time clustering of the different NPIs being applied typically in close sequence. NPIs largely reduce *R*_*t*_ by reducing contact among individuals. An indirect measure of the reduction in individual contact is the mobility data, and so these data will be highly correlated with NPIs, making any inference difficult by default. Moreover, as different NPIs are typically introduced in close sequence, their exact time lag before impact is difficult to model. Interaction effects between different NPIs may also exist. The effectiveness of different NPIs may also vary across locations and across time based on adherence, acceptability, and enforcement. Any collateral harms may also affect acceptability and adherence.

Given that the inference around the effectiveness of various NPIs is highly model dependent and that more aggressive NPIs have more adverse effects on other aspects of health, society, and economy ^7;8;9;10;11;12;13;14;15;16;17;18;19;20;21;22;23;24;25^, it is ill-advised to ignore the substantial model uncertainty. Failing to report this uncertainty may ultimately undermine the public’s trust in the value of policy decisions based on statistical modeling. Flaxman et al. ^1^ made the statement “We find that, across 11 countries, since the beginning of the epidemic, 3,100,000 [2,800,000 - 3,500,000] deaths have been averted due to intervention”. Both the provided estimate and the accompanying limited uncertainty are highly misleading. When results vary widely based on model specification, strong inferences should be avoided.

We are concerned that Flaxman et al. ^1^ selectively reported on only model 1, even though the Google mobility data was available from early April and the Imperial College team had obviously been using this data and both models 1 and 2, as evidenced by several of their reports^2;26;27^, before their *Nature* publication. The results included in the *Nature* paper seem to suffer from serious selective reporting, providing the most favorable estimates for lockdown benefits, while model 2 would have led to more nuanced, if not different, conclusions. Also the three European countries excluded from the Nature publication had among the least favorable results for lockdown.

Given that modeling studies are typically not pre-registered, multiple analytical approaches and model specifications may be used on the same data^28^, and data and results may be filtered by modelers according to whether they fit their prior beliefs. This bias can have devastating implication if it leads to adoption of harmful measures.

We do not claim that lockdown measures definitely had no impact in the first wave of COVID-19. Indeed model 2 showed that *R*_*t*_ was still above 1 in some countries and thus it is possible that in these locations it may have some impact on the course of the epidemic wave. Other investigators using a different analytical approach have suggested also some benefits from lockdown; however, these benefits were of a smaller magnitude (e.g. 13% relative risk reduction^29^). Small benefits of such modest size would be less likely to match complete lockdown-induced harms in a careful decision analysis. Another modeling approach has found that benefits can be reaped by simple self-imposed interventions such as washing hands, wearing masks, and some social distancing^30^.

Some limitations of our work should be acknowledged. Besides model fit and parsimony metrics, theoretical and subjective considerations, as well as experience from other countries should be considered in model choice. However, given the observational nature of the data and the dynamic course of epidemic waves, one should avoid strong priors about effectiveness of different NPIs. Similarly, our results should not be interpreted with a nihilistic lens, i.e. that NPIs are totally ineffective. Decreasing exposures makes sense as a way to reduce epidemic wave propagation and eventually fatalities. However, if exposures can be reduced with less aggressive measures and fewer or no harms, this would be optimal. Finally, we did not examine very long-term time horizons. In theory, even effective measures may achieve only temporary mitigation and epidemic waves may surge again, when measures are relieved. We did observe this for the uplifting of measures in the July 12th analyses and empirical data from the emergence of second waves in many European countries and the USA in the fall of 2020 validate this hypothesis^31^. Availability of effective and safe vaccines may also affect risk-benefit ratios of NPI measures of different aggressiveness and different duration of implementation.

Overall, observational data that feed into complex epidemic models should be dissected very carefully and substantial uncertainty may remain despite the best efforts of modelers^28;32^. While there has been resistance to testing NPIs with randomized trials, such trials are feasible, and more thought and effort should be devoted on how to complement the available, tenuous observational data^33^. Regardless, causal interpretations from non-robust models should be avoided. In any decision analysis the accurate quantification of the size, not just the existence, of the impact of lockdown on *R*_*t*_ is also critical. This is difficult task when one considers all the confounds between NPIs and mobility, as well as the several behavioral changes such as hand washing and wearing masks. This is an interesting area for research, and crucial for the management of future pandemics.

## Data Availability

All source code for the replication of our results is available from https://github.com/dare-centre/imperial-covid19-model

## Acknowledgements

We congratulate the Imperial College Response Team for sharing openly the code for their models and for the overall transparency of their work that has allowed performing these analyses. We thank Hadi Ashfar for his suggestions to improve the computational efficiency of the HMC scheme. We also thank Jack Wood for his help in the construction of Table A.3. We acknowledge the Sydney Informatics Hub and the University of Sydney’s high performance computing cluster Artemis for providing the high performance computing resources that have contributed to the research results reported within this paper.

## Author Contributions

All authors contributed equally to this work. VC performed all the computations and produced all the graphics. SC wrote the initial draft. JI and MT wrote subsequent drafts. All authors discussed the results and implications and commented on the manuscript at all stages.

## A. Appendix: Additional Figures and Tables

**Figure A.1:**
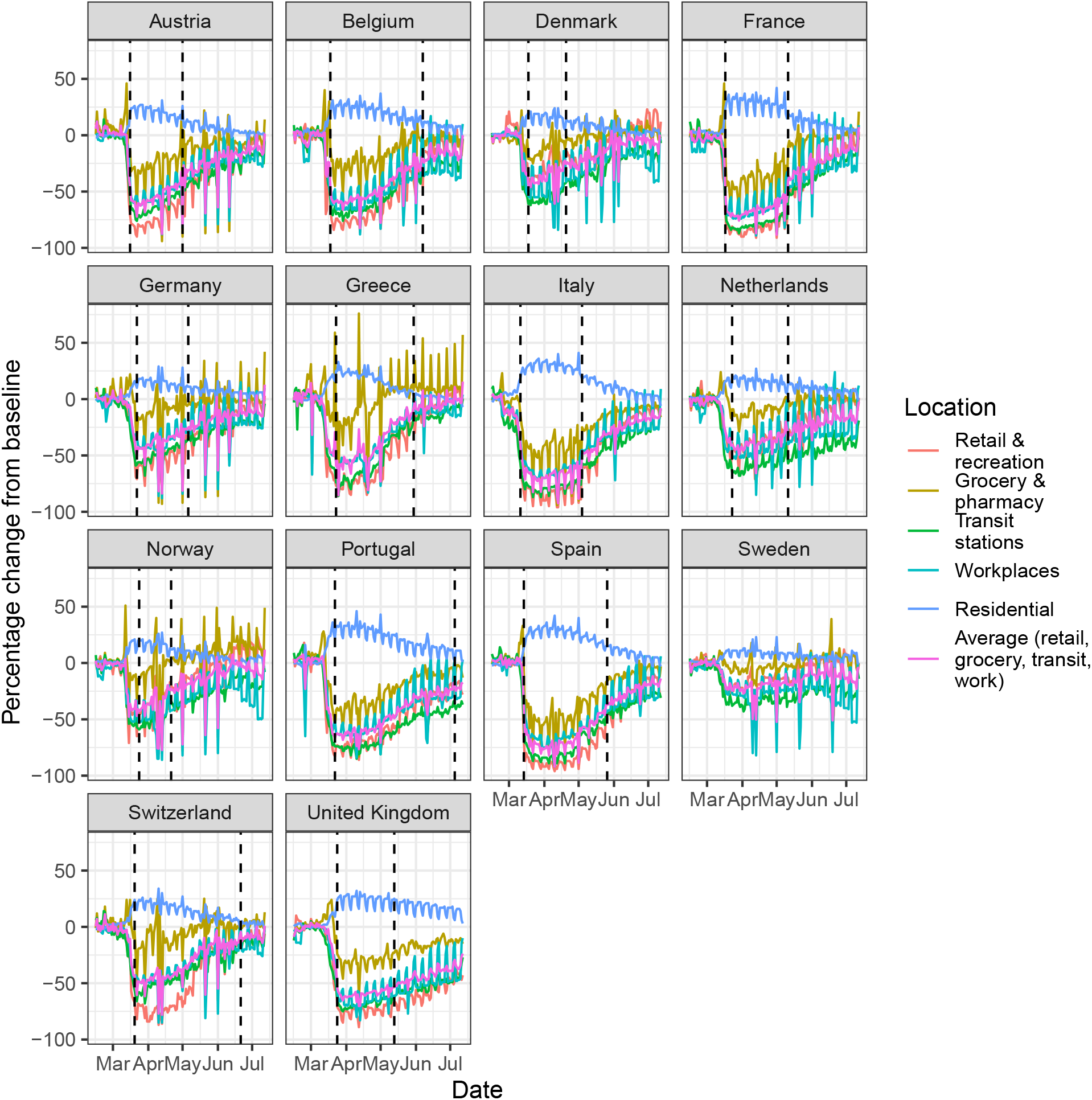
Percentage change in mobility from baseline level from February 15th to July 12th, by locations in each of the European countries examined in Flaxman et al. ^1^, as well as an additional three countries consisting of Greece, the Netherlands and Portugal. Average mobility is computed based on the trends in retailers and recreation venues, grocery markets and pharmacy, transit stations and workplaces. Black dashed lines in each plot indicate the lockdown start and end dates.

**Figure A.2:**
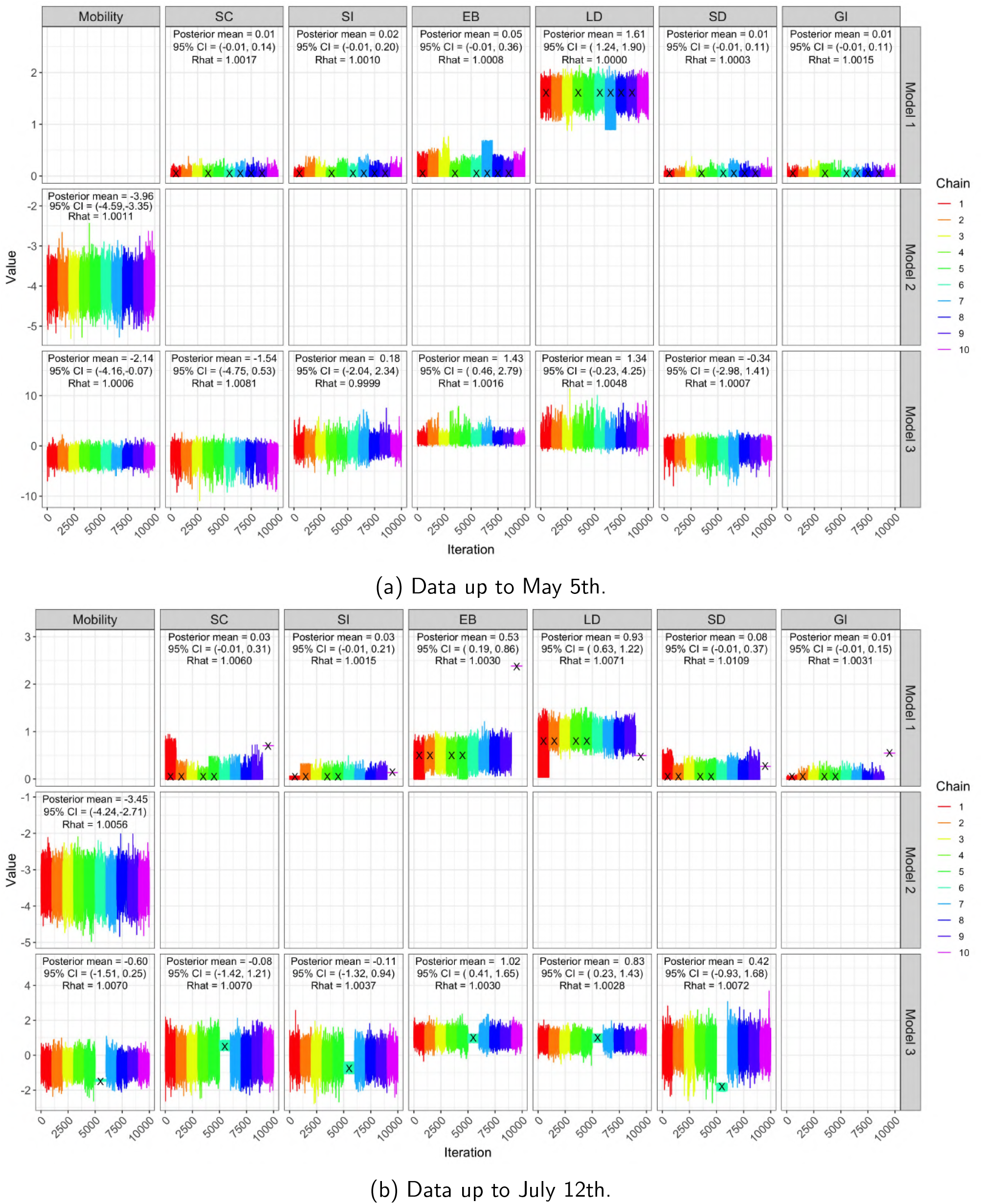
Trace plots, posterior mean, 95% CIs and 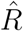 of regression coefficients in *R*_*t*_ for all three models based on ten MCMC chains, for both time horizons. All our analyses use the iterates from chains which had no more than 5% of divergent transitions. Black crosses (×) denote chains with more than 5% of divergent transitions.

**Figure A.3:**
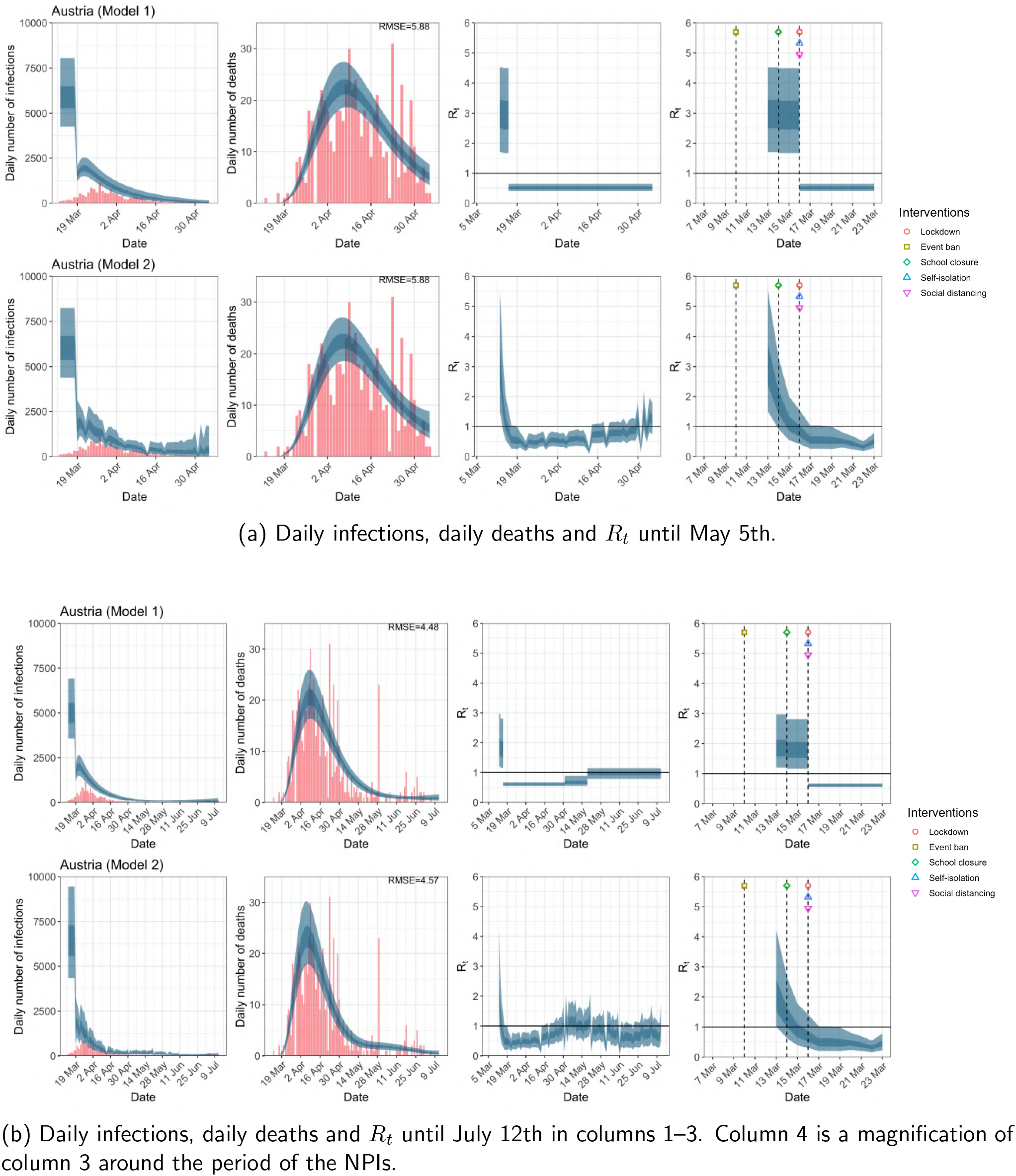
Austria. The start time for the plots is 10 days before 10 deaths are recorded. Observed counts of daily infections and daily deaths are shown in red, and their corresponding 50% and 95% CIs are shown in dark blue and light blue respectively.

**Figure A.4:**
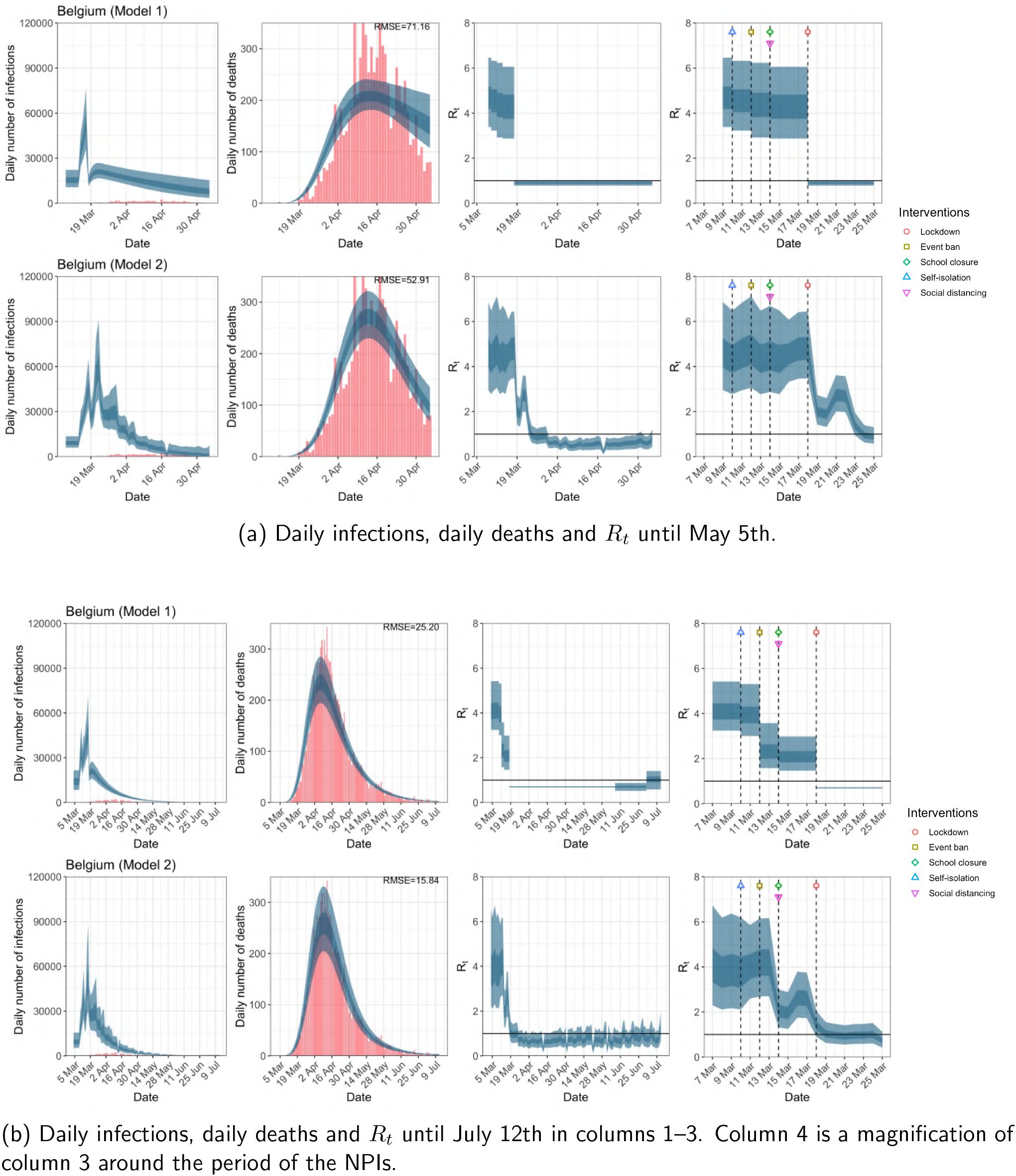
Belgium. The start time for the plots is 10 days before 10 deaths are recorded. Observed counts of daily infections and daily deaths are shown in red, and their corresponding 50% and 95% CIs are shown in dark blue and light blue respectively.

**Figure A.5:**
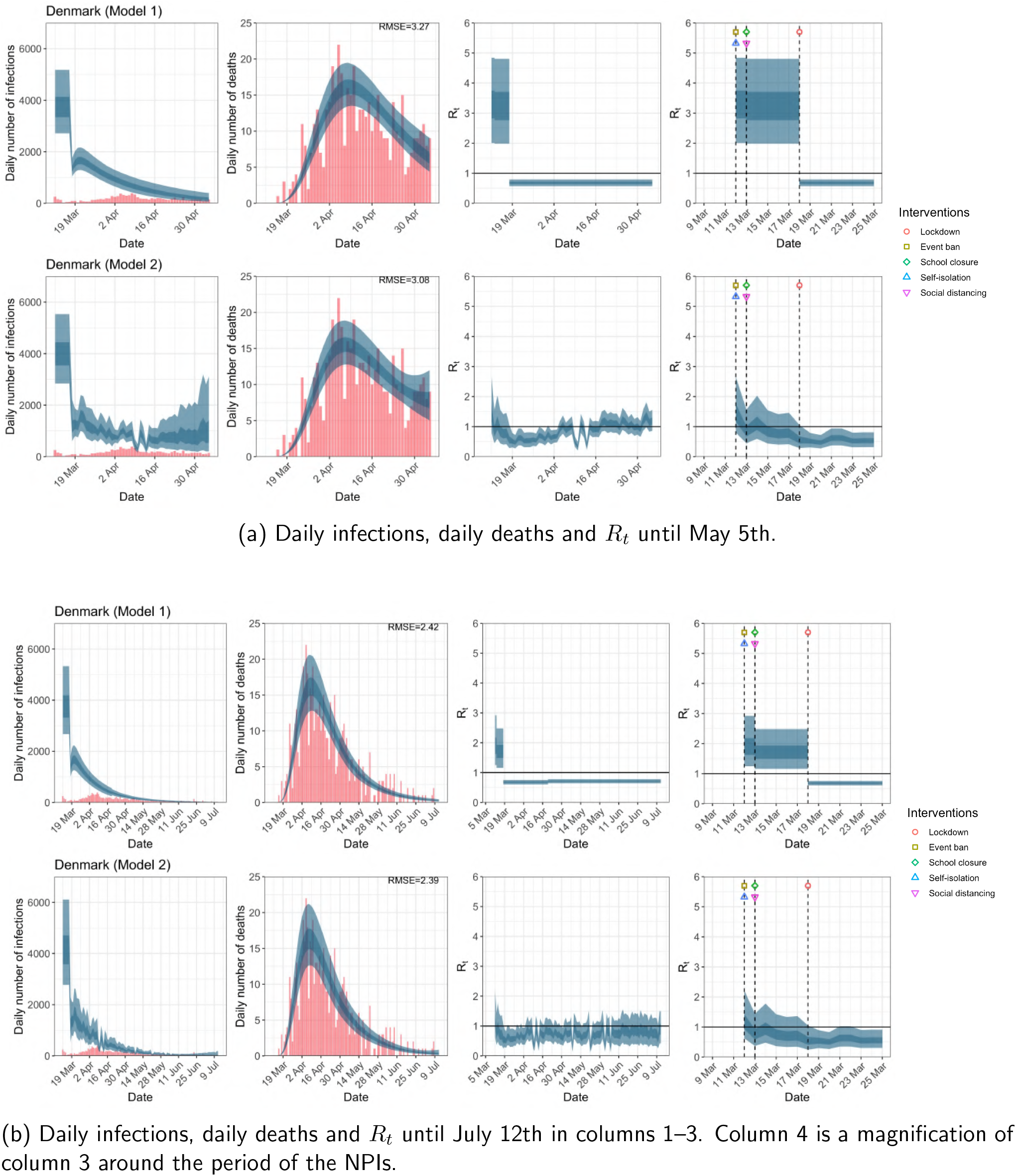
Denmark. The start time for the plots is 10 days before 10 deaths are recorded. Observed counts of daily infections and daily deaths are shown in red, and their corresponding 50% and 95% CIs are shown in dark blue and light blue respectively.

**Figure A.6:**
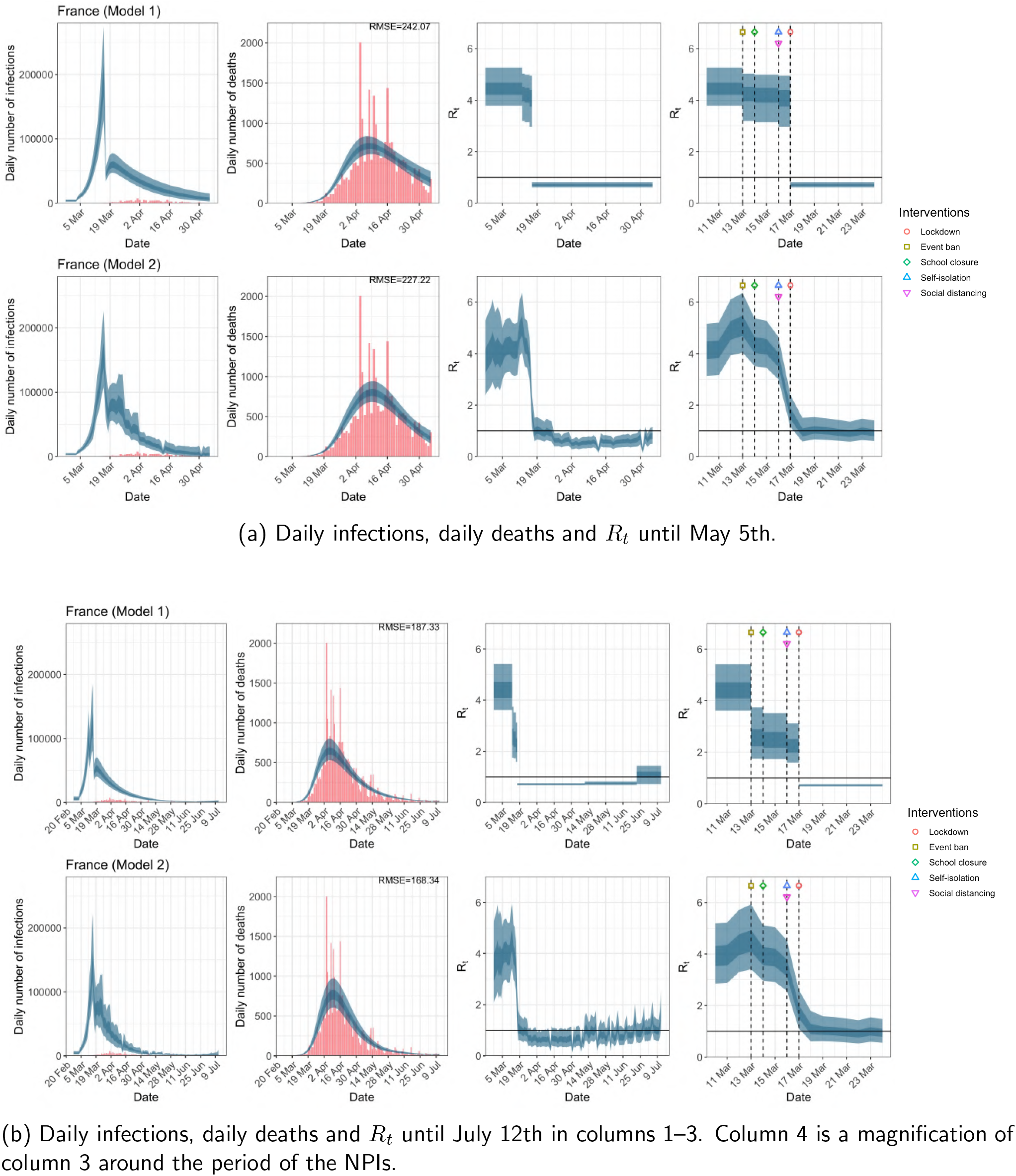
France. The start time for the plots is 10 days before 10 deaths are recorded. Observed counts of daily infections and daily deaths are shown in red, and their corresponding 50% and 95% CIs are shown in dark blue and light blue respectively.

**Figure A.7:**
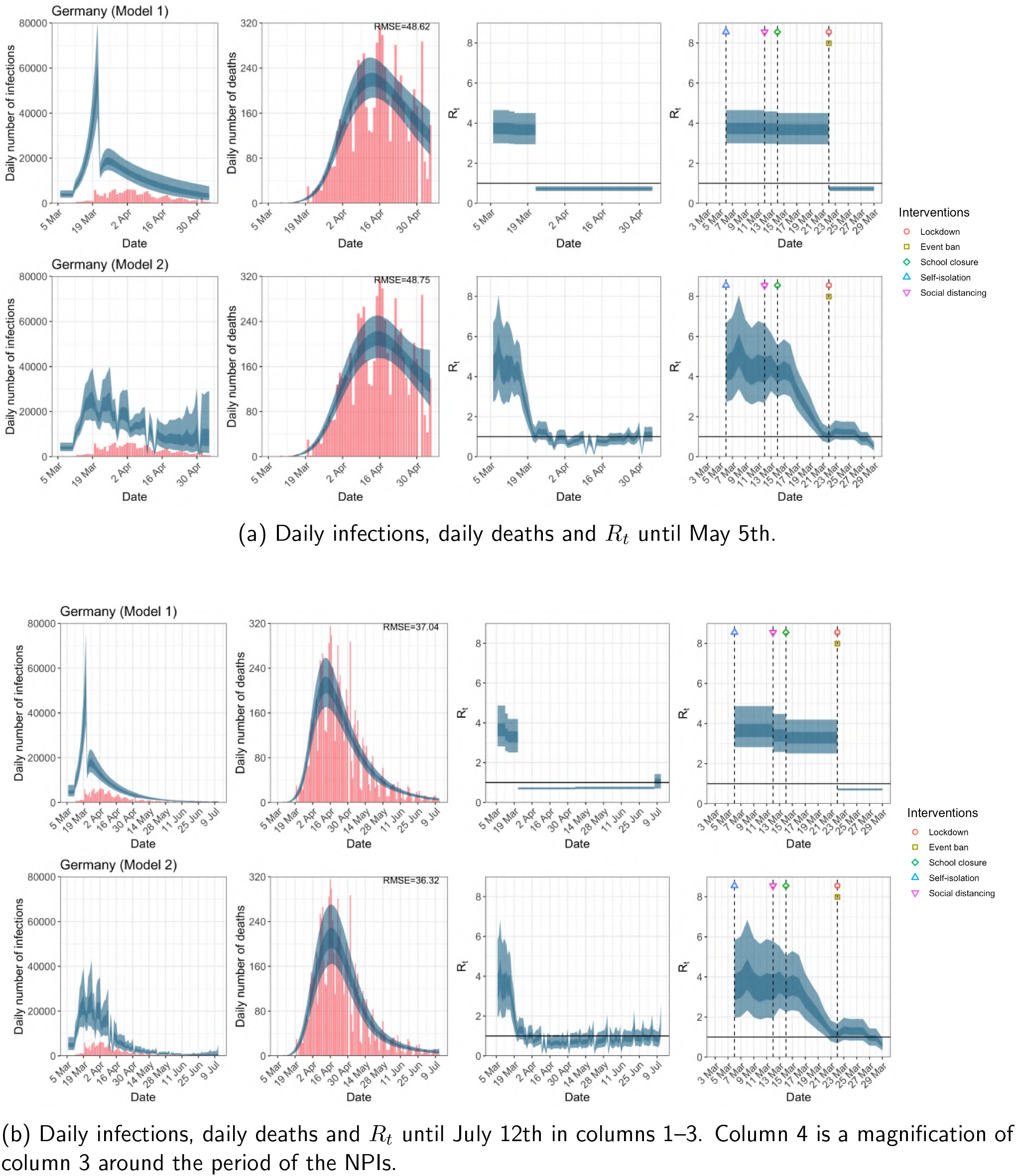
Germany. The start time for the plots is 10 days before 10 deaths are recorded. Observed counts of daily infections and daily deaths are shown in red, and their corresponding 50% and 95% CIs are shown in dark blue and light blue respectively.

**Figure A.8:**
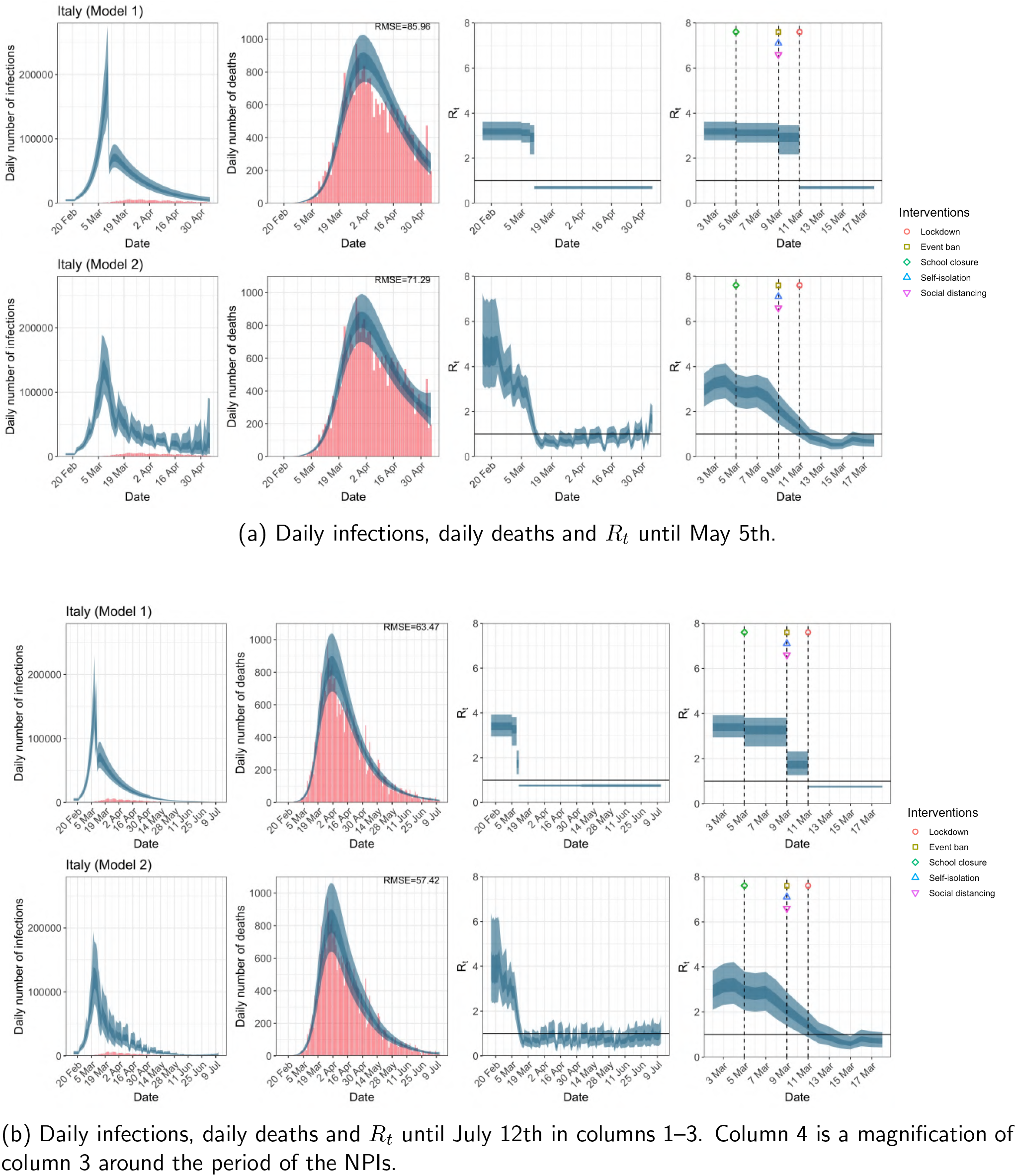
Italy. The start time for the plots is 10 days before 10 deaths are recorded. Observed counts of daily infections and daily deaths are shown in red, and their corresponding 50% and 95% CIs are shown in dark blue and light blue respectively.

**Figure A.9:**
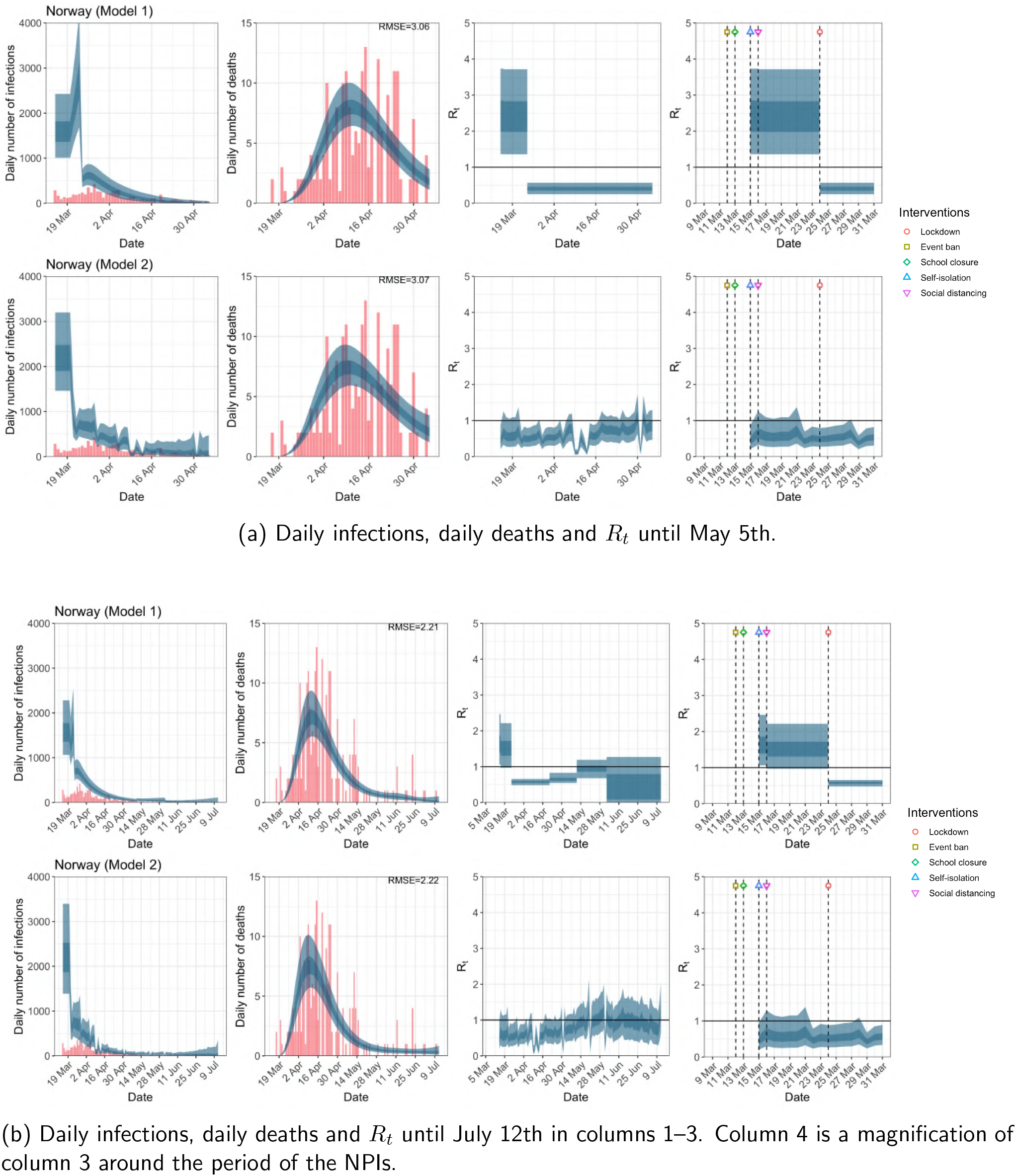
Norway. The start time for the plots is 10 days before 10 deaths are recorded. Observed counts of daily infections and daily deaths are shown in red, and their corresponding 50% and 95% CIs are shown in dark blue and light blue respectively.

**Figure A.10:**
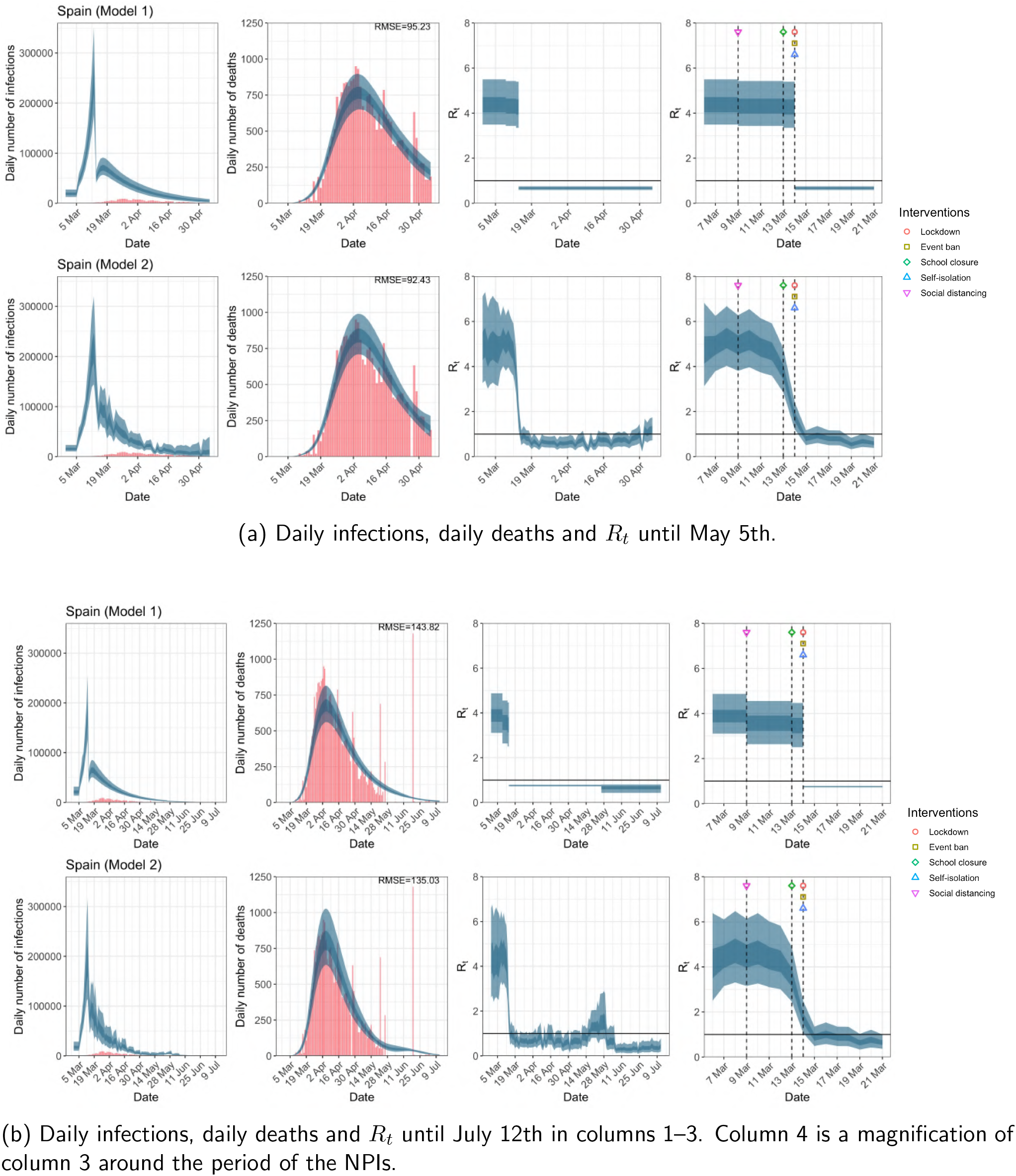
Spain. The start time for the plots is 10 days before 10 deaths are recorded. Observed counts of daily infections and daily deaths are shown in red, and their corresponding 50% and 95% CIs are shown in dark blue and light blue respectively.

**Figure A.11:**
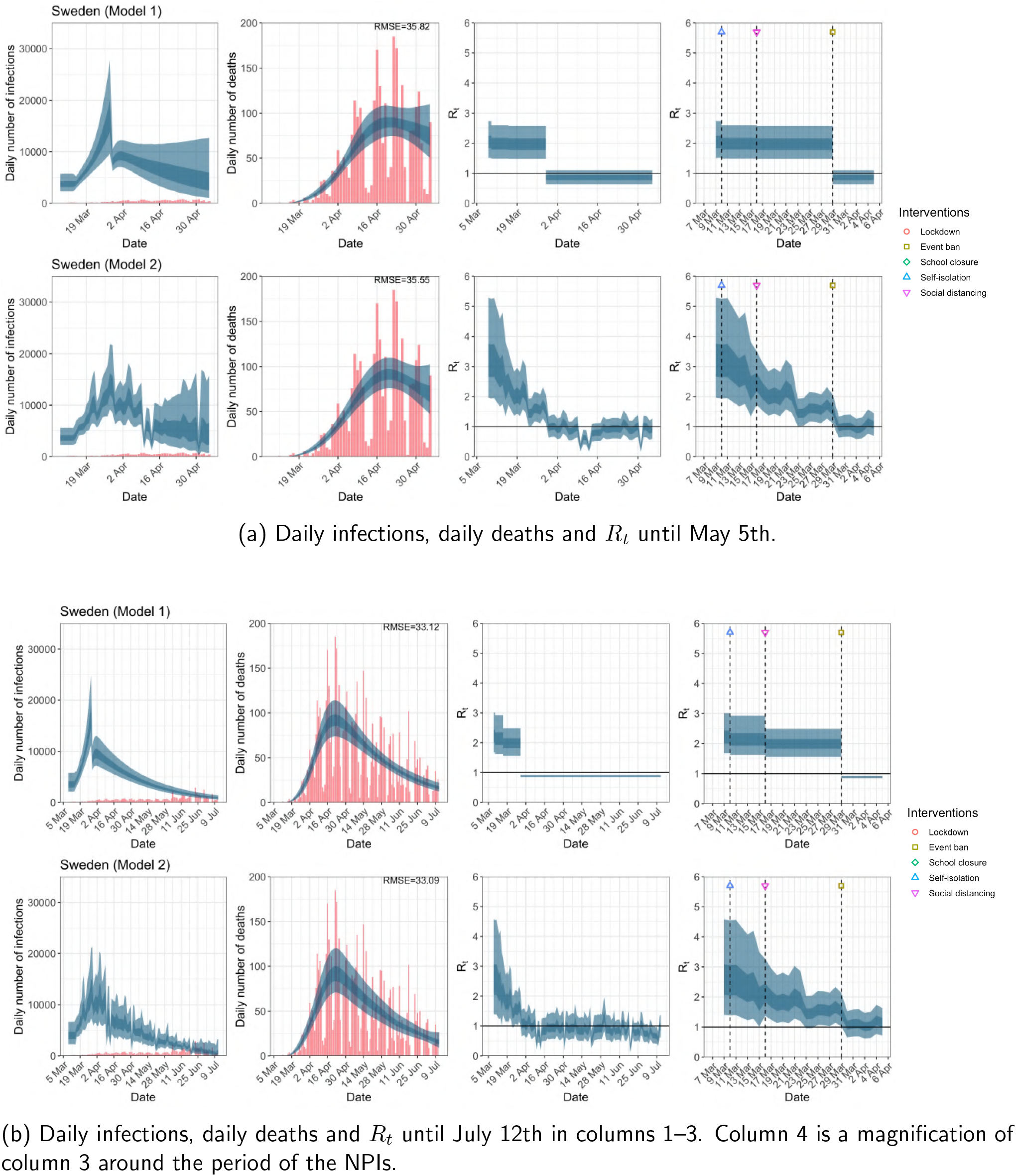
Sweden. The start time for the plots is 10 days before 10 deaths are recorded. Observed counts of daily infections and daily deaths are shown in red, and their corresponding 50% and 95% CIs are shown in dark blue and light blue respectively.

**Figure A.12:**
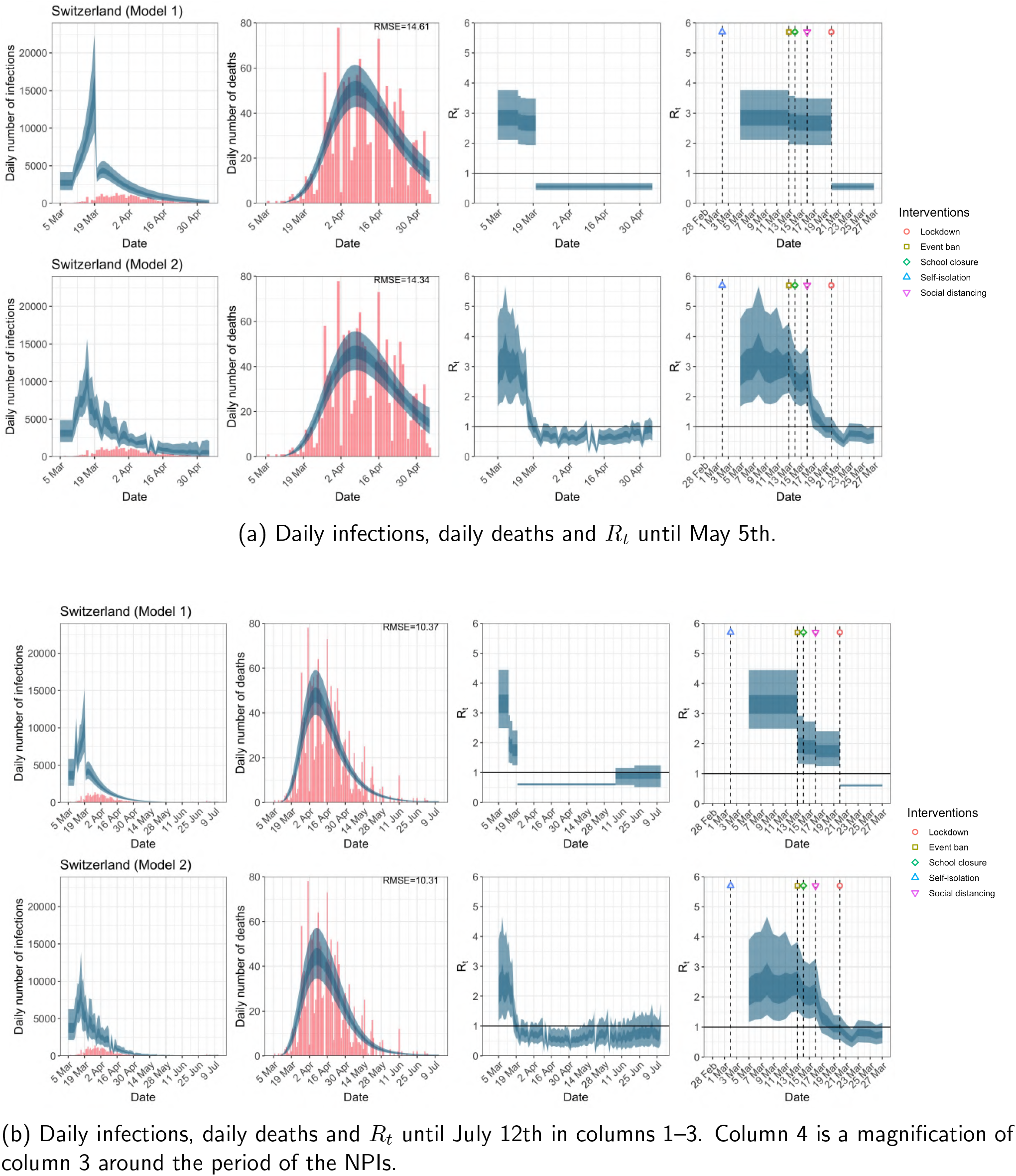
Switzerland. The start time for the plots is 10 days before 10 deaths are recorded. Observed counts of daily infections and daily deaths are shown in red, and their corresponding 50% and 95% CIs are shown in dark blue and light blue respectively.

**Figure A.13:**
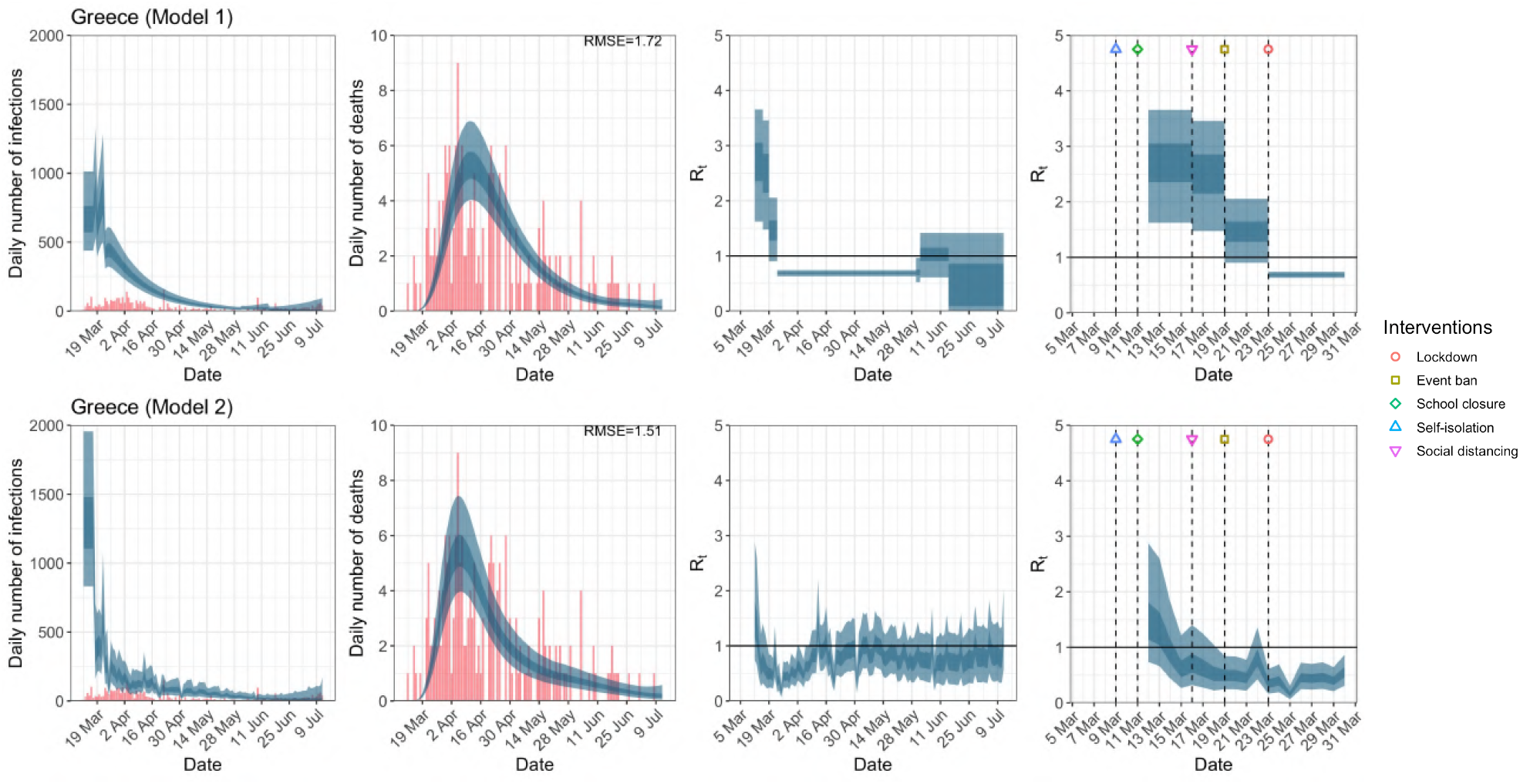
Greece. The start time for the plots is 10 days before 10 deaths are recorded. Observed counts of daily infections and daily deaths until July 12th are shown in red, and their corresponding 50% and 95% CIs are shown in dark blue and light blue respectively. Column 4 is a magnification of column 3 showing the changes in *R*_*t*_ around the period of the NPIs.

**Figure A.14:**
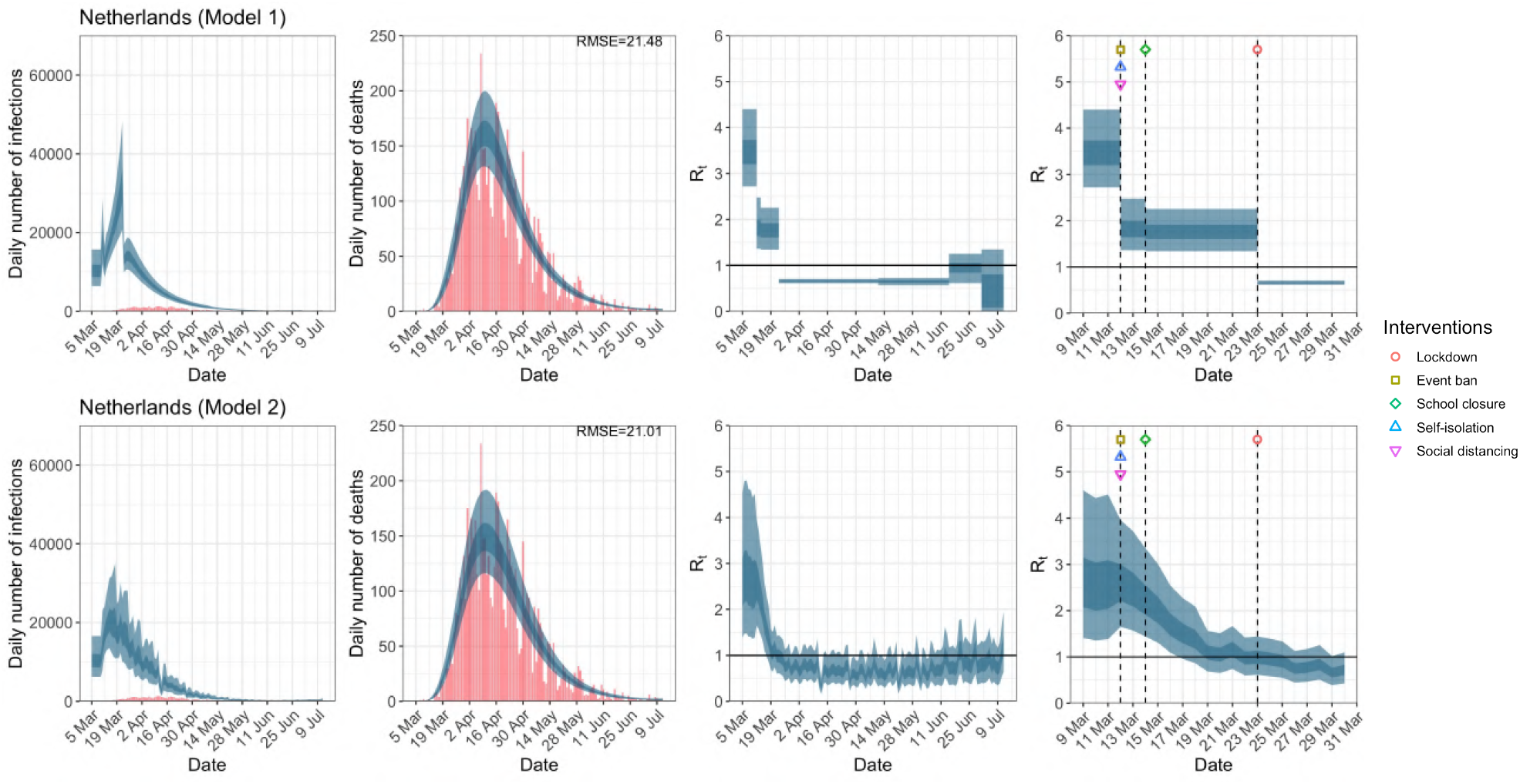
Netherlands. The start time for the plots is 10 days before 10 deaths are recorded. Observed counts of daily infections and daily deaths until July 12th are shown in red, and their corresponding 50% and 95% CIs are shown in dark blue and light blue respectively. Column 4 is a magnification of column 3 showing the changes in *R*_*t*_ around the period of the NPIs.

**Figure A.15:**
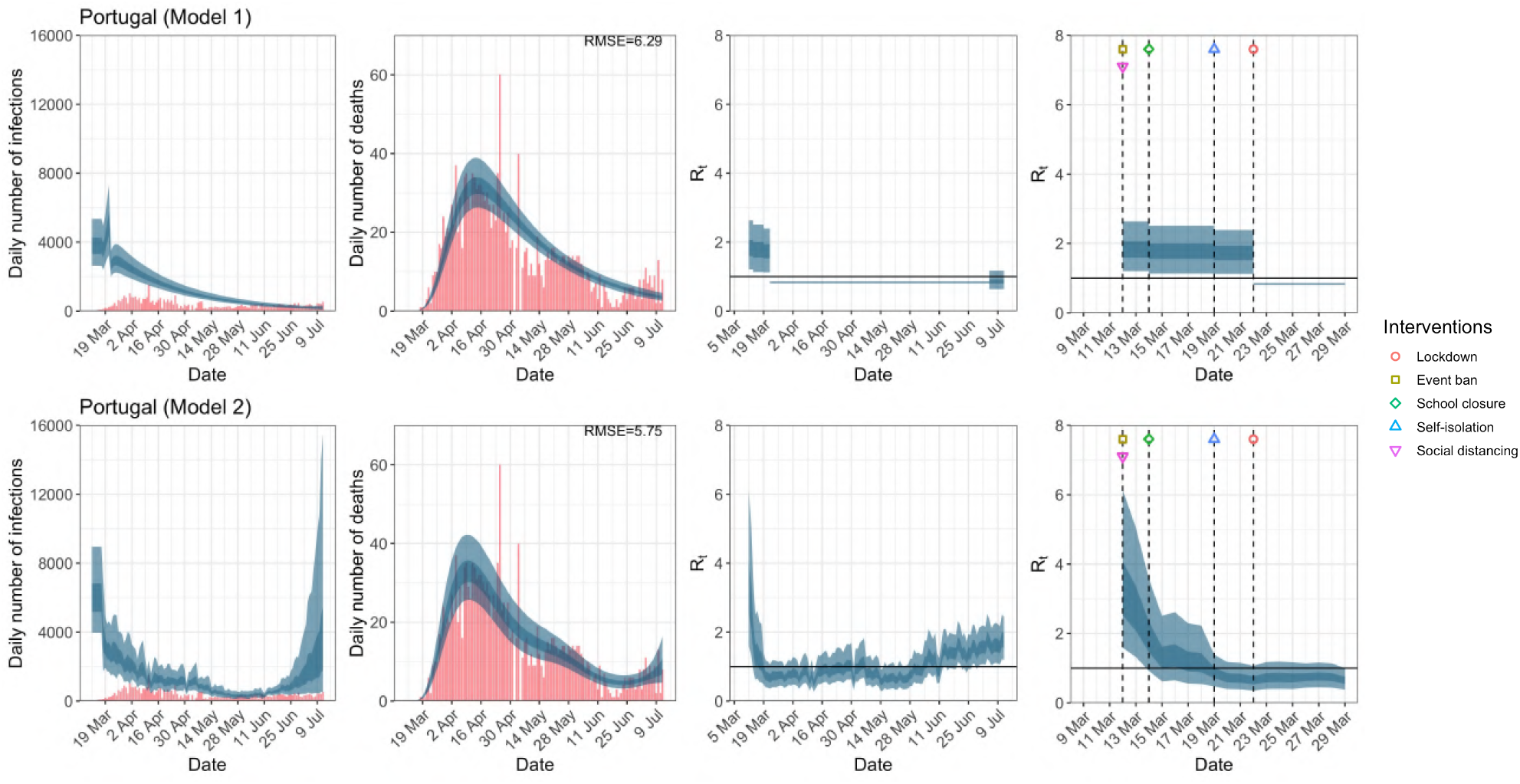
Portugal. The start time for the plots is 10 days before 10 deaths are recorded. Observed counts of daily infections and daily deaths until July 12th are shown in red, and their corresponding 50% and 95% CIs are shown in dark blue and light blue respectively. Column 4 is a magnification of column 3 showing the changes in *R*_*t*_ around the period of the NPIs.

**Figure A.16:**
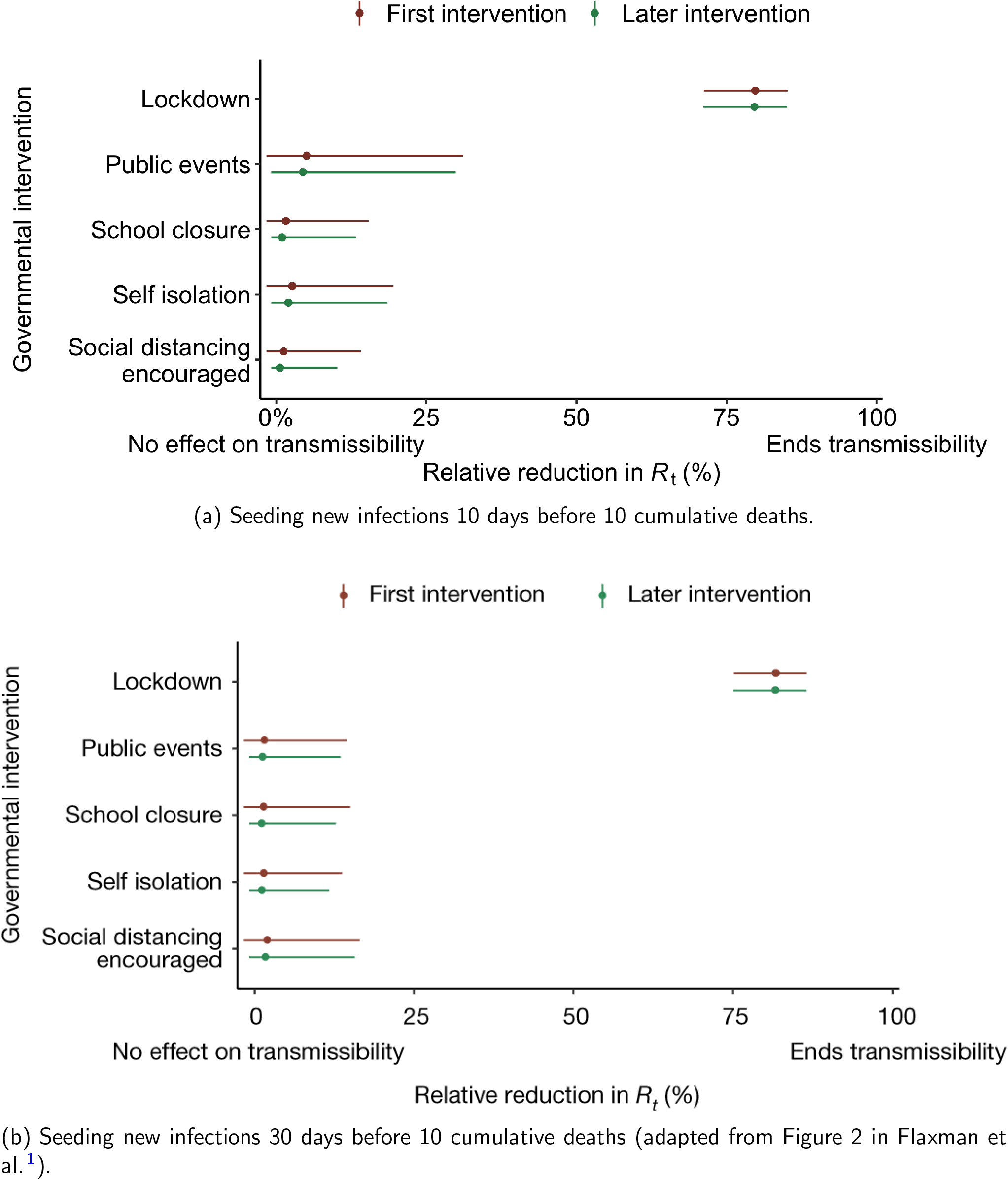
Comparison of the effectiveness of NPIs in terms of the relative percentage reduction in *R*_*t*_ when assuming two different seeding periods of new infections.

**Table A.1:** Seeding dates of new infections. Two seeding dates were used for Belgium – March 9th and March 4th for the data up to May 5th and July 12th respectively due to a reporting correction in the data.

| Country | Seeding date |
| --- | --- |
| Austria | March 13th |
| Belgium | March 9th / March 4th |
| Denmark | March 12th |
| France | February 27th |
| Germany | March 6th |
| Greece | March 12th |
| Italy | February 16th |
| Netherlands | March 5th |
| Norway | March 15th |
| Portugal | March 12th |
| Spain | February 29th |
| Sweden | March 9th |
| Switzerland | March 5th |

**Table A.2:** Correspondence of subscripts for *k* to each NPI.

| $k$ | NPIs |
| --- | --- |
| 1 | school closure |
| 2 | event ban |
| 3 | lockdown |
| 4 | self-isolation |
| 5 | social distancing |
| 6 | government intervention |

**Table A.3:** End dates for school closure^34^, event ban^34^ and lockdown in each country^35;36;37^. NPIs that are still in place as of July 12th are shown in ✓, while NPIs that were not implemented are shown in ✗.

| Country | School closure | Event ban | Lockdown |
| --- | --- | --- | --- |
| UK | ✓ | ✓ | May 13th |
| Austria | May 18th | ✓ | May 1st |
| Belgium | July 1st | ✓ | June 7th |
| Denmark | ✓ | ✓ | April 20th |
| France | June 22th | ✓ | May 11th |
| Germany | July 7th | ✓ | May 6th |
| Greece | June 1st | June 15th | May 30th |
| Italy | ✓ | ✓ | May 4th |
| Netherlands | June 15th | July 1st | May 11th |
| Norway | May 11th | June 2nd | April 21st |
| Portugal | ✓ | ✓ | July 5th |
| Spain | ✓ | ✓ | May 26th |
| Sweden | ✗ | ✓ | ✗ |
| Switzerland | June 6th | ✓ | June 21st |

**Table A.4:** RMSE of daily death counts for model 1 and 2 for the data up to May 5th and July 12th.

| Country | Up to May 5th |  | Up to July 12th |  |
| --- | --- | --- | --- | --- |
|  | Model 1 | Model 2 | Model 1 | Model 2 |
| UK | **145.41** | 145.64 | 134.26 | **129.68** |
| Austria | **5.88** | **5.88** | **4.48** | 4.57 |
| Belgium | 71.16 | **52.91** | 25.20 | **15.84** |
| Denmark | 3.27 | **3.08** | 2.42 | **2.39** |
| France | 242.07 | **227.22** | 187.33 | **168.34** |
| Germany | **48.62** | 48.75 | 37.04 | **36.32** |
| Italy | 85.96 | **71.29** | 63.47 | **57.42** |
| Norway | **3.06** | 3.07 | **2.21** | 2.22 |
| Spain | 95.23 | **92.43** | 143.82 | **135.03** |
| Sweden | 35.82 | **35.55** | 33.12 | **33.09** |
| Switzerland | 14.61 | **14.34** | 10.37 | **10.31** |
| Greece |  |  | 1.72 | **1.51** |
| Netherlands |  |  | 21.48 | **21.01** |
| Portugal |  |  | 6.29 | **5.75** |

**Table A.5:**
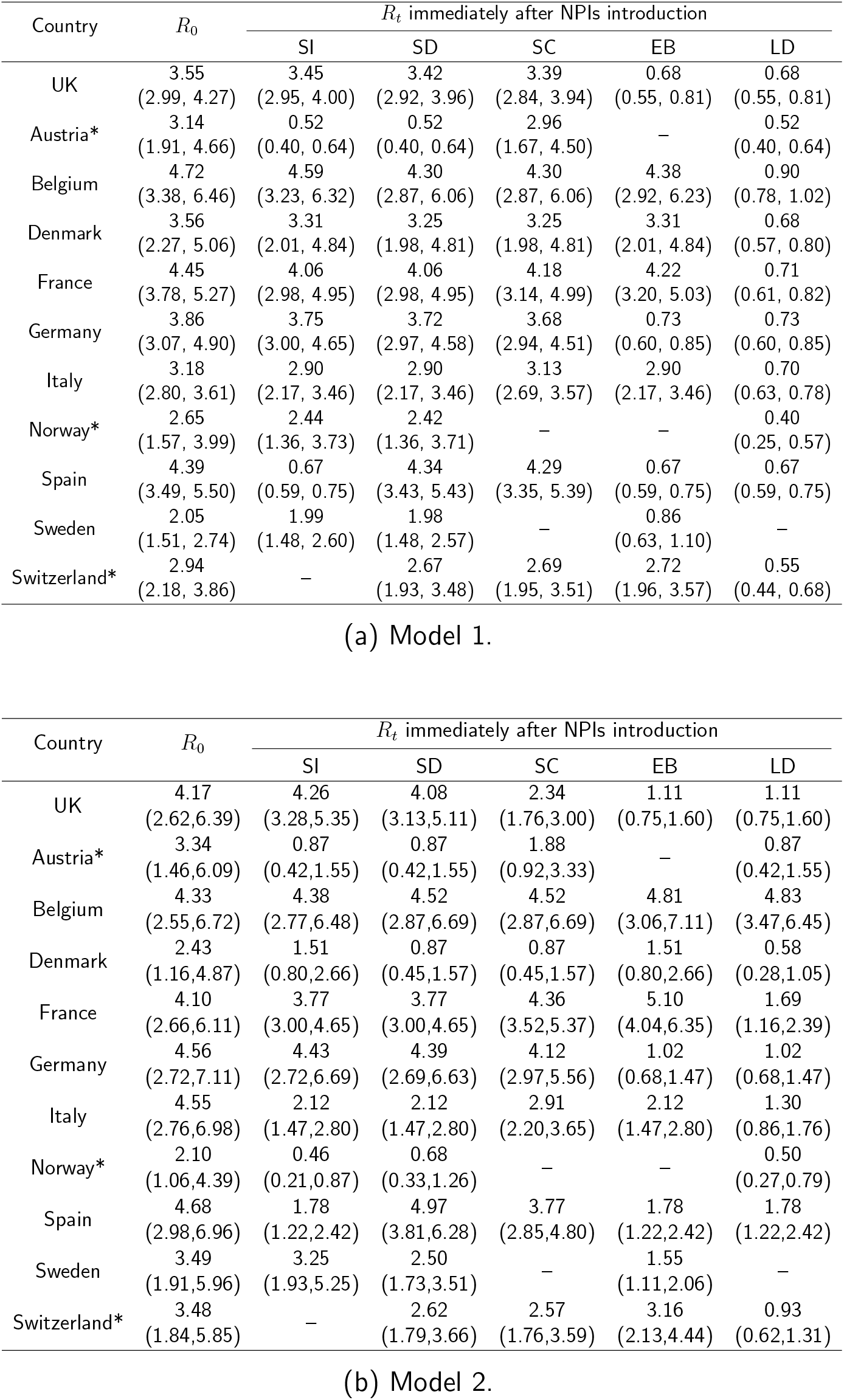
Basic reproduction number *R*_0_ and time-varying reproduction number *R*_*t*_ immediately after the introduction of NPIs given by both models using data up to May 5th for all eleven countries analysed in Flaxman et al.^1^. These NPIs are self-isolation (SI), social distancing (SD), school closure (SC), event ban (EB) and lockdown (LD). 95% credible intervals are given in parentheses below the corresponding point estimates. Countries where the seeding of new infections occur after the introduction of NPIs are denoted with an asterisk.

## B. Appendix: Priors and Measures of Fit

### B.1 Priors

For posterior inference in model 1, we use the same priors as in Flaxman et al.^1^ for the analysis up to May 5th and July 12th. For model 2, we use the same prior distributions as in Unwin et al.^2^ except for *R*_0_, and *α* in Equation 2.

For *R*_0_ we use a weakly informative prior of a normal distribution truncated below at 1 with mean 3.28 and standard deviation 2 is used. This prior is chosen so that approximately 95% of the prior density is between 1 and 7^38^, and that *R*_0_ is above the critical value of 1 at the start of the epidemic.

For *α*, we examine the sensitivity of the posterior to two priors. The first prior that we consider is that used by Unwin et al.^2^–this prior is very informative, with *α* ∼*N* (0, 0.5). That is, *a priori* they assume *α* lies in the interval [−1, 1] with probability 0.95. In contrast, the second prior we considered was an uninformative prior, *α* ∼*N* (0, 5), and found the posterior mode of *α* in model 2 up to May 5th to be approximately −4. This means that the prior used by Unwin et al.^2^ has almost no support over this posterior distribution. This has two consequences, first it makes convergence of the Markov chain very difficult and sensitive to starting values. Second, it shrinks the value of *α* towards zero, underestimating the impact of mobility on *R*_*t*_. The second prior, *α* ∼*N* (0, 5), makes the convergence of the chain more robust to poor starting values.

We also change the prior for the number of initial infection at the start of the time period for two reasons. First, due to data constraints, we chose to start the seeding of infections only 10 days before the date of the 10th cumulative death. In contrast, Flaxman et al. ^1^ chose to start the seeding of infections 30 days prior to the date of the 10th cumulative death. Flaxman et al. ^1^ chose a prior for initial infection count which was relatively tight, the probability that the initial infection count was greater than 500 is ≈ 0. Using this prior for the number of infections 20 days later again, is not realistic and again leads to convergence problems. We therefore chose a less informative prior for the initial infection count. Plots of these prior distributions can be found in Figure B.1.

Notably, Bayesian data analysts typically examine a variety of priors to gauge the sensitivity of results to the prior specification^6^. Flaxman et al^1^ limited analyses to one prior distribution for their entire set of parameters.

### B.2 Bayesian measures of model fit

To compare the fit of the three models to the data we consider four metrics; three estimates of various information criteria, as well as the root mean square error (RMSE). The information criteria metrics are two versions of the Watanabe-Akaike information criteria^39^, denoted by WAIC1 and WAIC2 and the Deviance information criterion DIC^40^. Both WAIC1 and WAIC2 use the log pointwise predictive density (lppd) as a measure of fit. To penalize the fit, WAIC1 uses

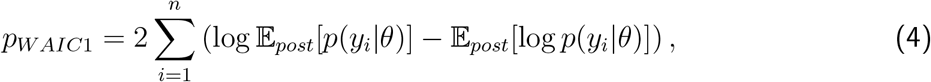

as an estimate of the effective number of parameters, where 𝔼_*post*_ denotes the expectation over the posterior distribution of model parameters *θ* given the observed data **y** = {*y*_*i*_; *i* = 1, …, *n*}.

**Figure B.1:**
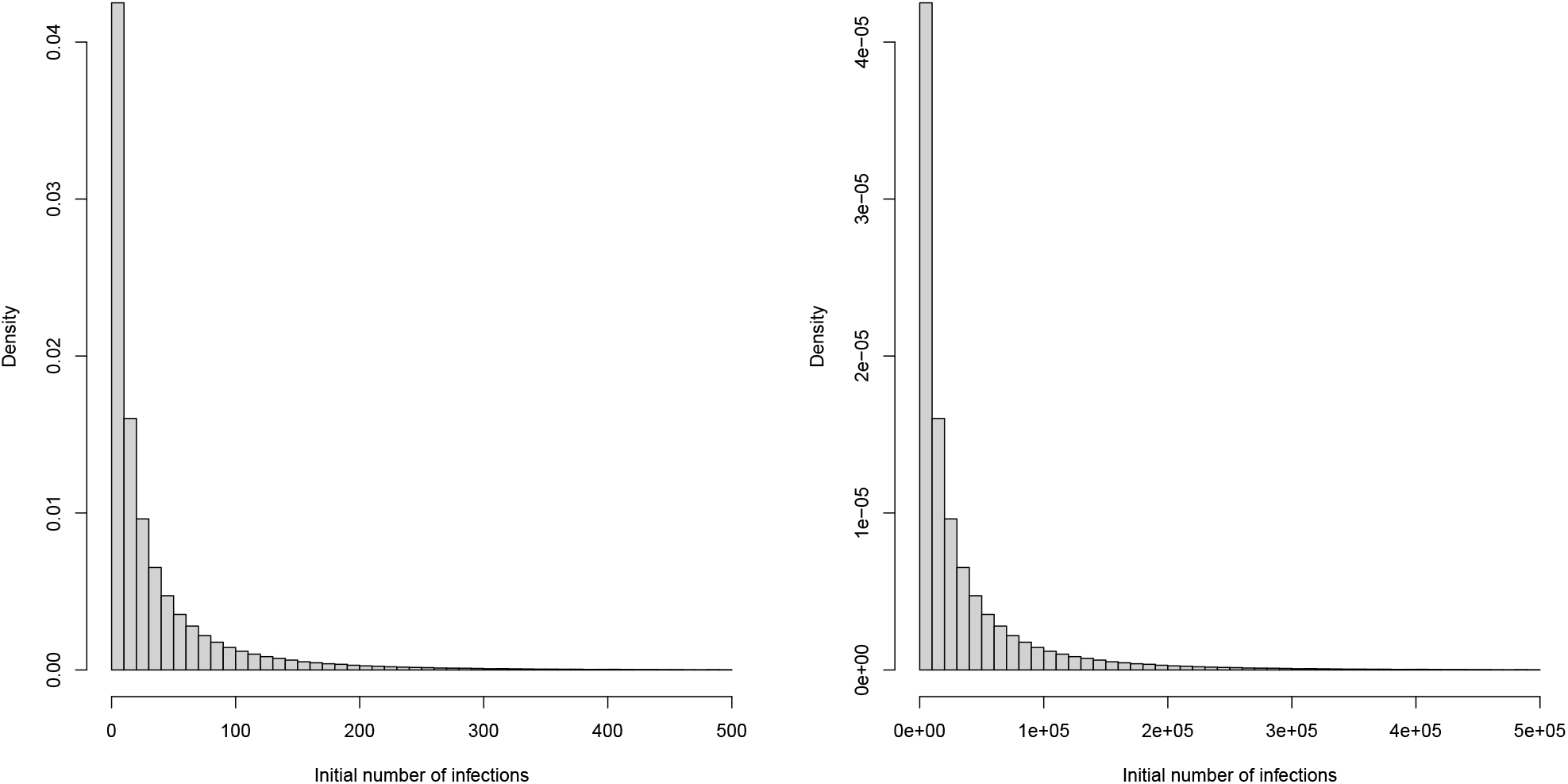
Prior distributions of initial infection count used in Flaxman et al. ^1^ (left) and our analysis (right).

The criteria WAIC2 uses

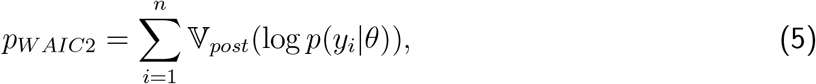

where 𝕍_*post*_ denotes the variance over the posterior distribution of *θ*. The DIC metric uses 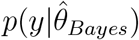, with 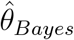 being the posterior mean of *θ*, as a measure of fit and

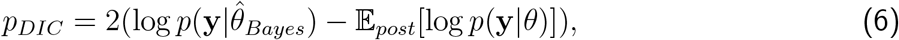

as the penalty.

## C. Appendix: Analysis up to May 5th for all 14 countries

**Figure C.1:**
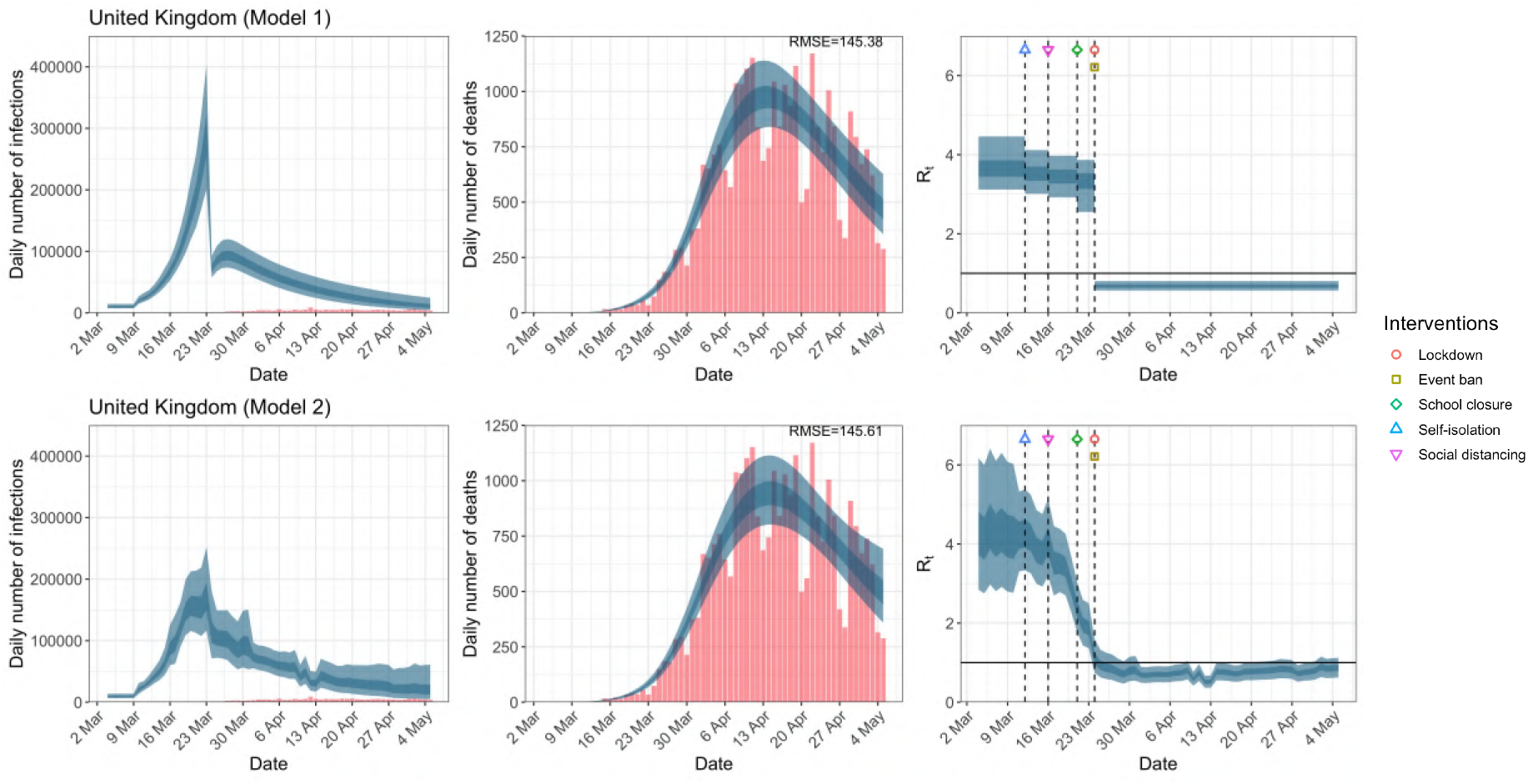
Daily infections, daily deaths and *R*_*t*_ in the United Kingdom until May 5th. The start time for the plots is 10 days before 10 deaths are recorded. Observed counts of daily infections and daily deaths are shown in red, and their corresponding 50% and 95% CIs are shown in dark blue and light blue respectively.

**Figure C.2:**
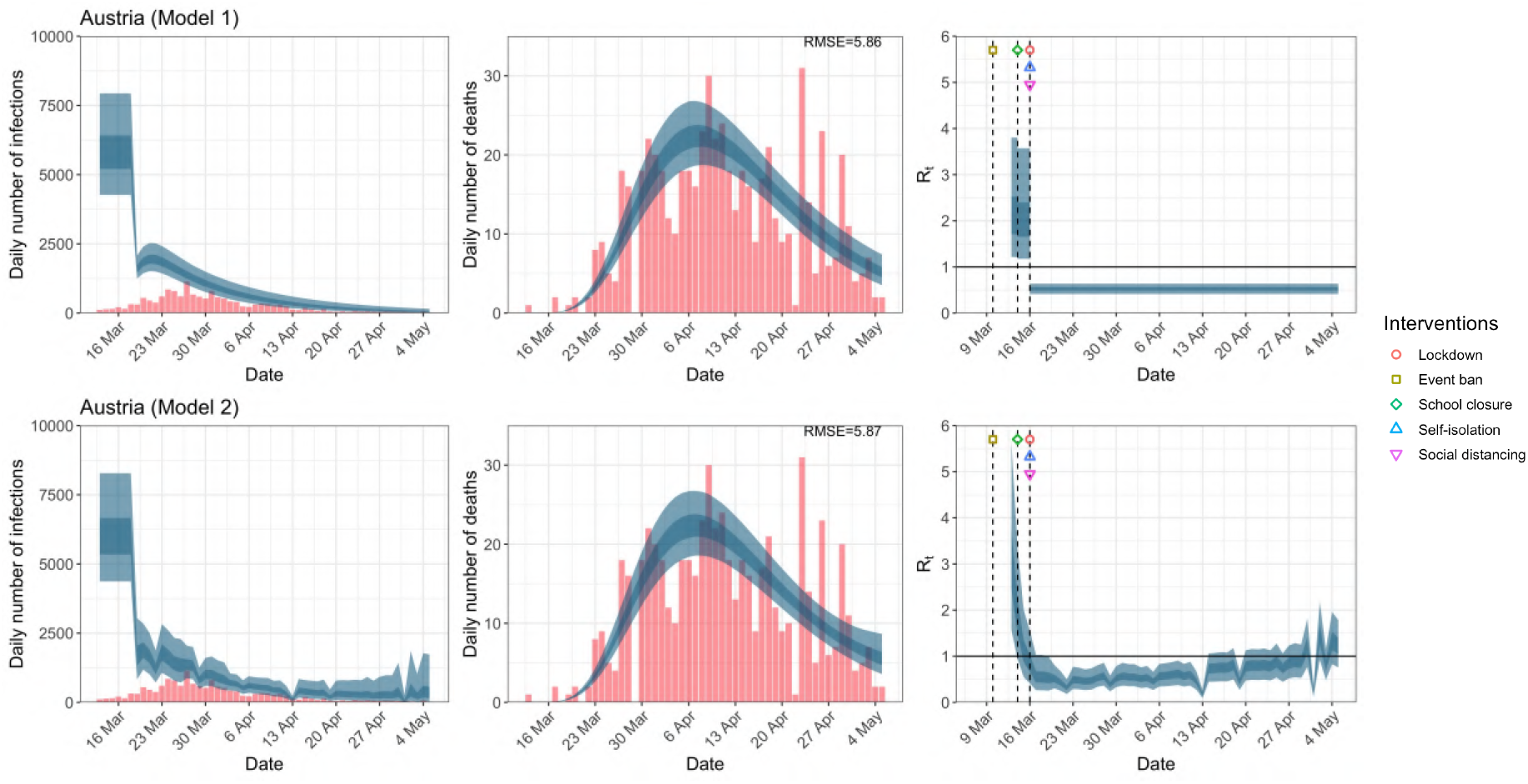
Daily infections, daily deaths and *R*_*t*_ in Austria until May 5th. The start time for the plots is 10 days before 10 deaths are recorded. Observed counts of daily infections and daily deaths are shown in red, and their corresponding 50% and 95% CIs are shown in dark blue and light blue respectively.

**Figure C.3:**
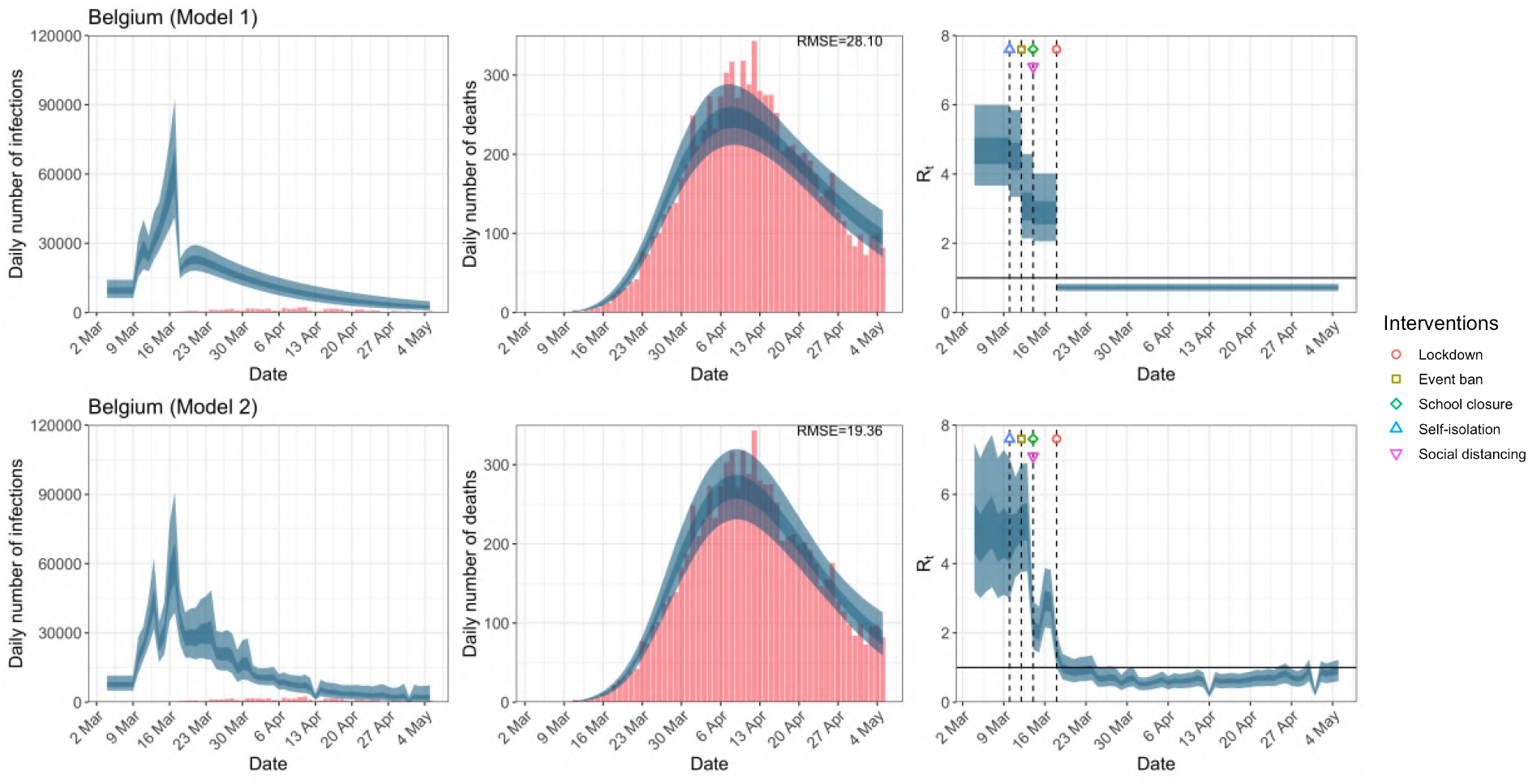
Daily infections, daily deaths and *R*_*t*_ in Belgium until May 5th. The start time for the plots is 10 days before 10 deaths are recorded. Observed counts of daily infections and daily deaths are shown in red, and their corresponding 50% and 95% CIs are shown in dark blue and light blue respectively.

**Figure C.4:**
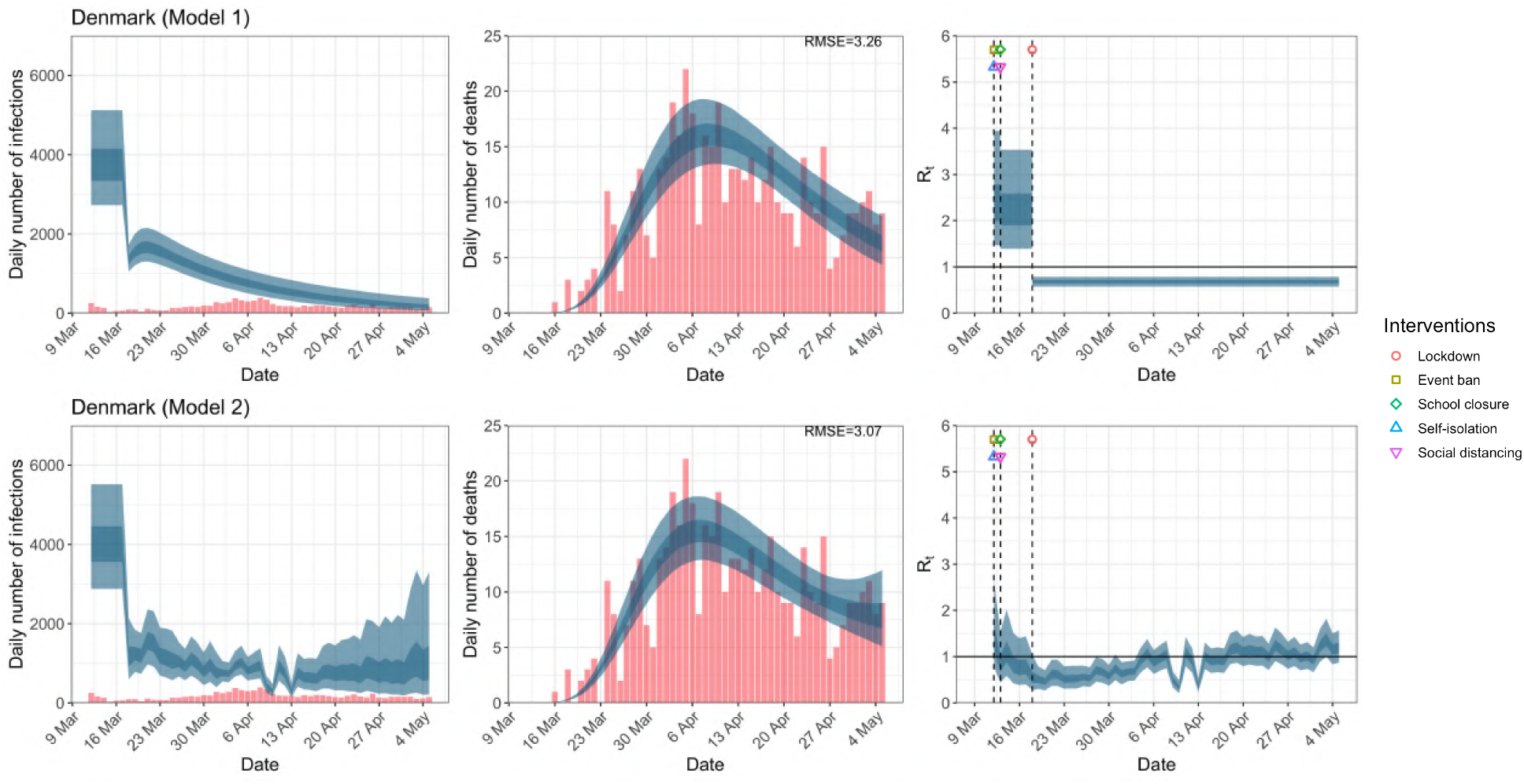
Daily infections, daily deaths and *R*_*t*_ in Denmark until May 5th. The start time for the plots is 10 days before 10 deaths are recorded. Observed counts of daily infections and daily deaths are shown in red, and their corresponding 50% and 95% CIs are shown in dark blue and light blue respectively.

**Figure C.5:**
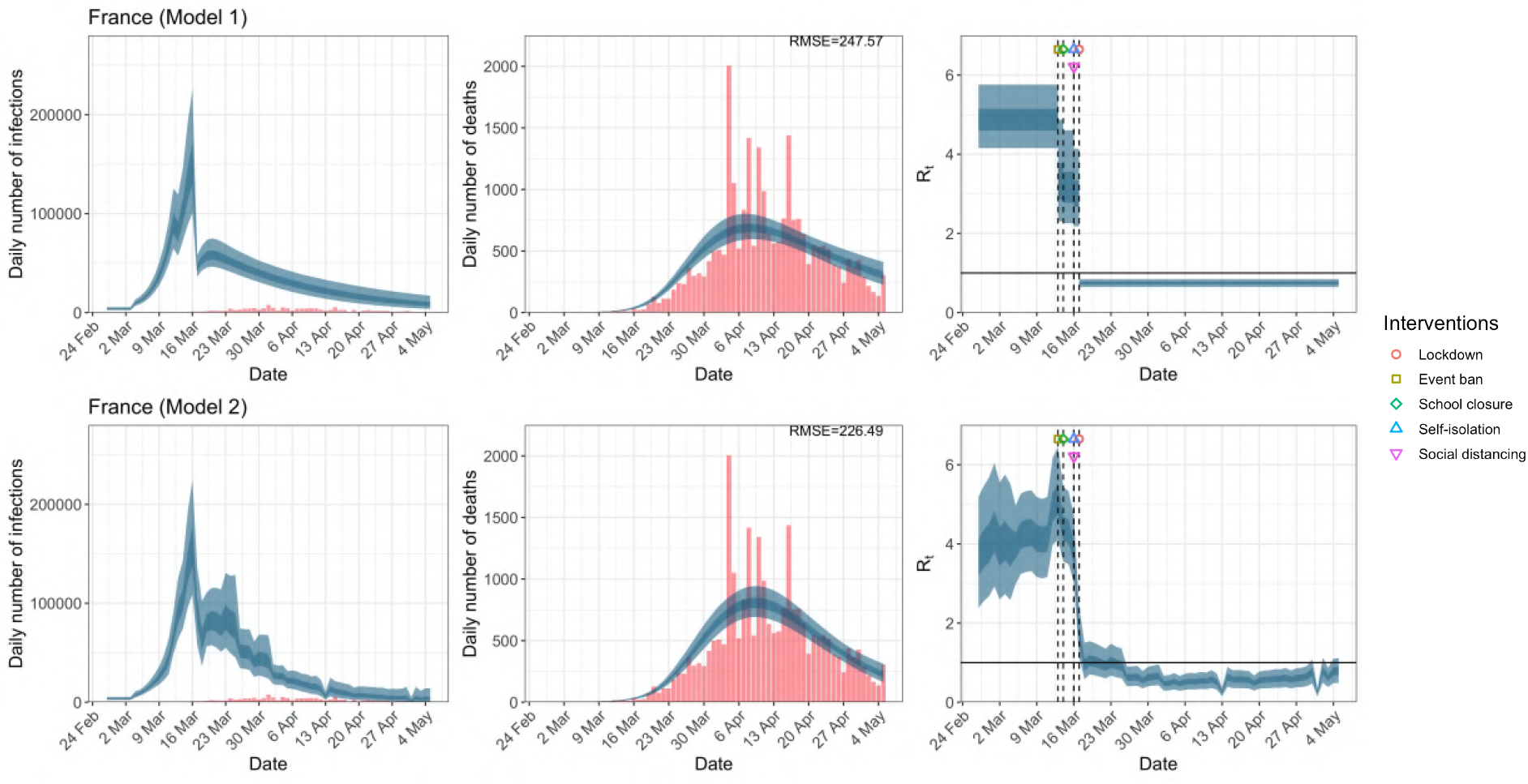
Daily infections, daily deaths and *R*_*t*_ in France until May 5th. The start time for the plots is 10 days before 10 deaths are recorded. Observed counts of daily infections and daily deaths are shown in red, and their corresponding 50% and 95% CIs are shown in dark blue and light blue respectively.

**Figure C.6:**
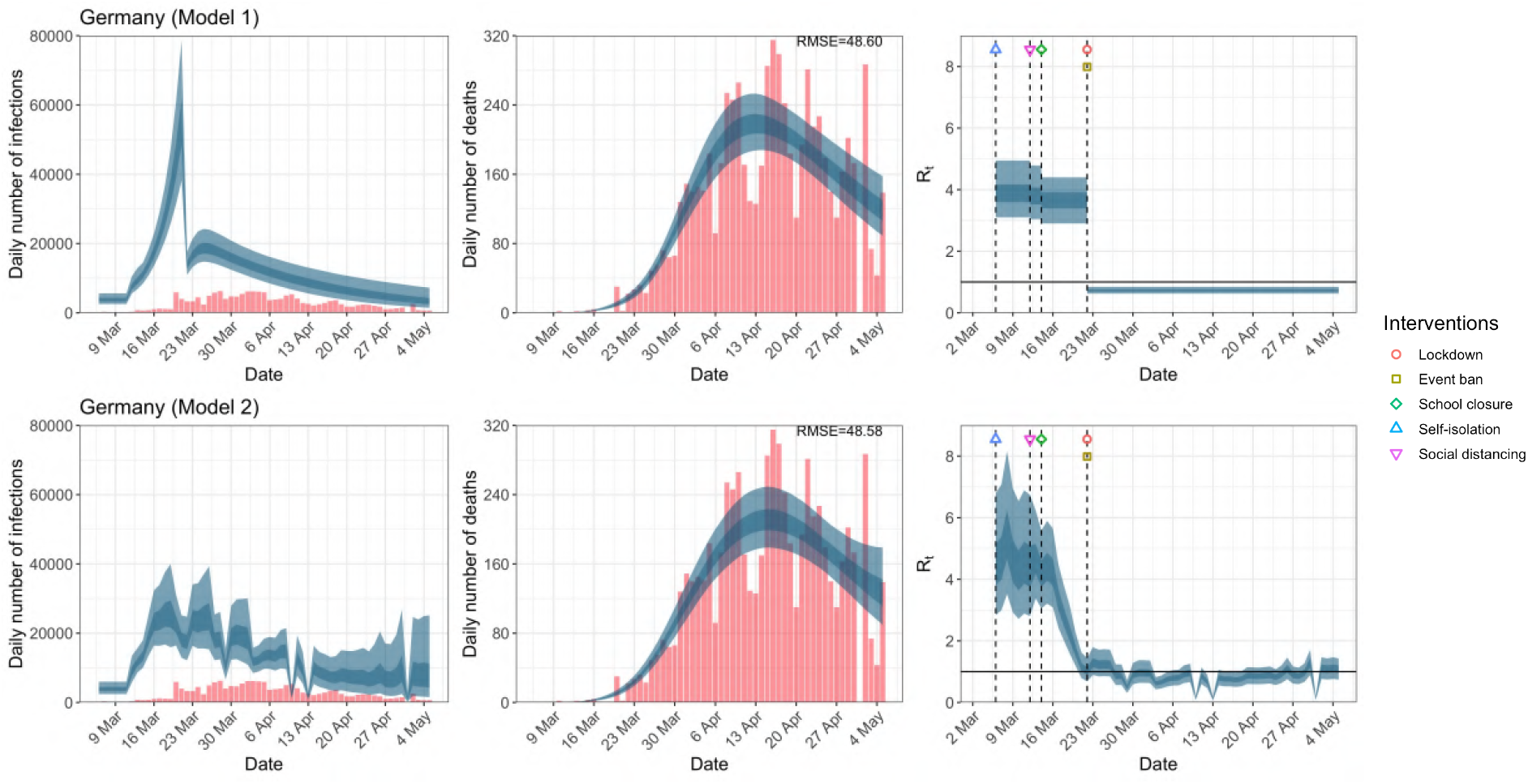
Daily infections, daily deaths and *R*_*t*_ in Germany until May 5th. The start time for the plots is 10 days before 10 deaths are recorded. Observed counts of daily infections and daily deaths are shown in red, and their corresponding 50% and 95% CIs are shown in dark blue and light blue respectively.

**Figure C.7:**
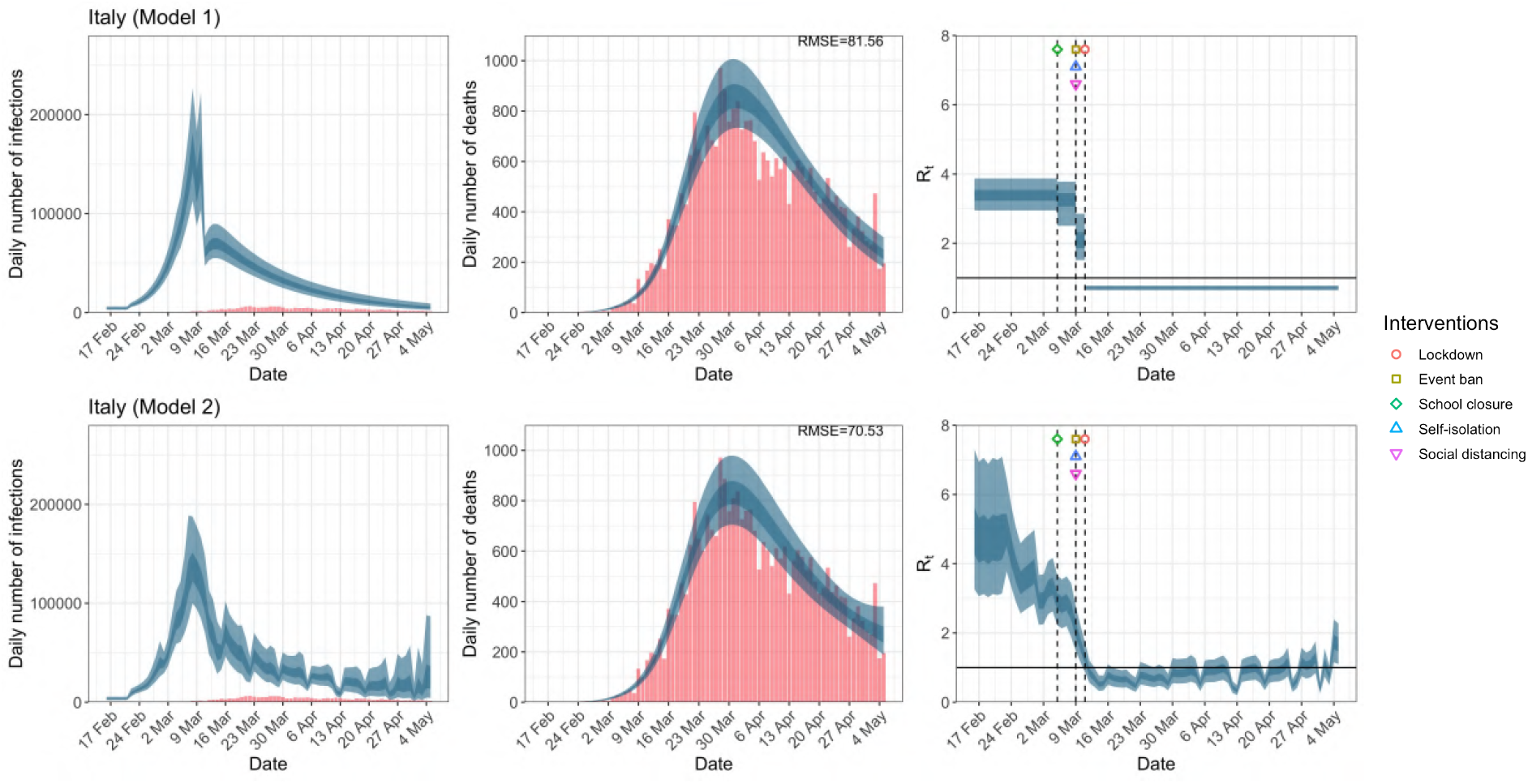
Daily infections, daily deaths and *R*_*t*_ in Italy until May 5th. The start time for the plots is 10 days before 10 deaths are recorded. Observed counts of daily infections and daily deaths are shown in red, and their corresponding 50% and 95% CIs are shown in dark blue and light blue respectively.

**Figure C.8:**
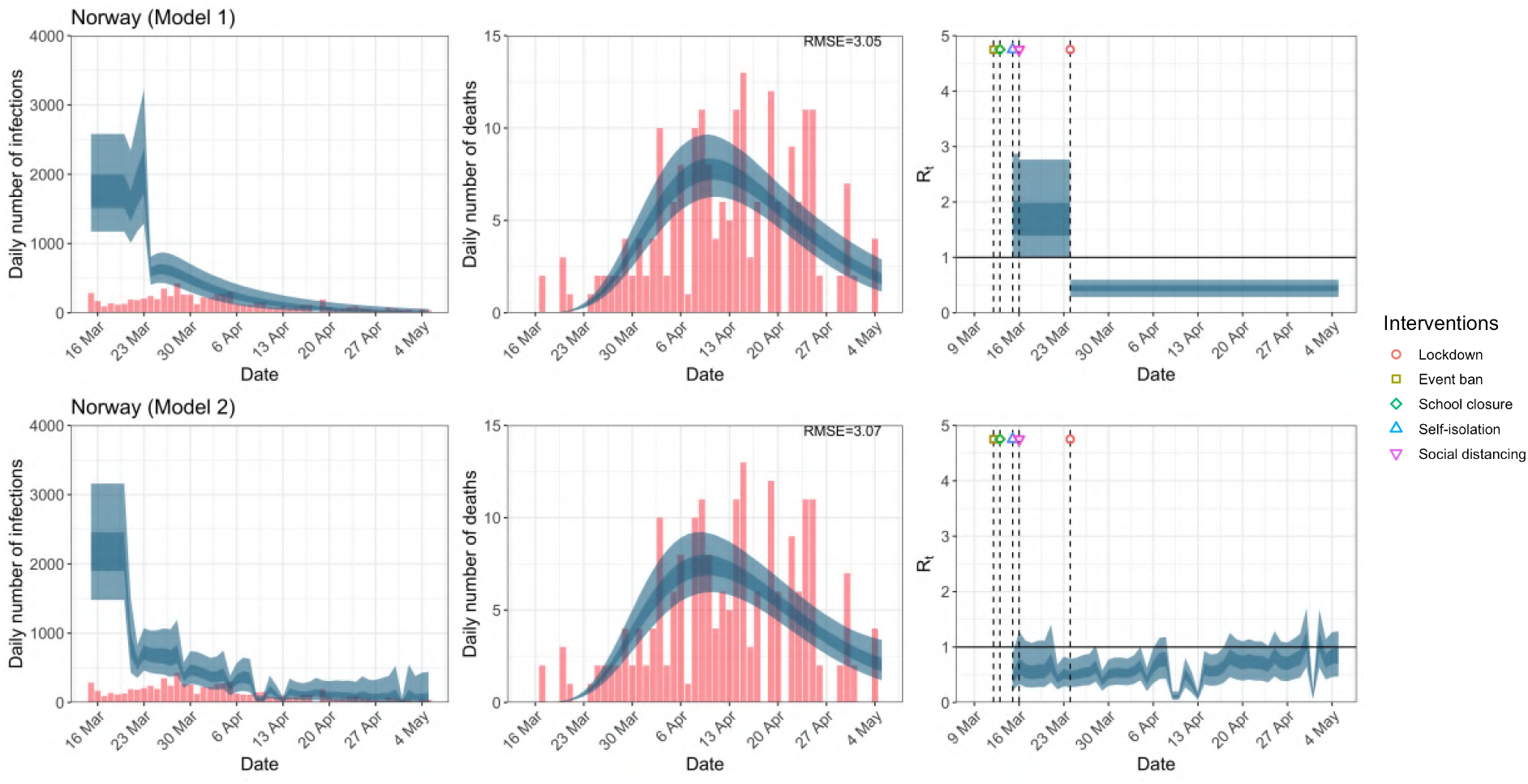
Daily infections, daily deaths and *R*_*t*_ in Norway until May 5th. The start time for the plots is 10 days before 10 deaths are recorded. Observed counts of daily infections and daily deaths are shown in red, and their corresponding 50% and 95% CIs are shown in dark blue and light blue respectively.

**Figure C.9:**
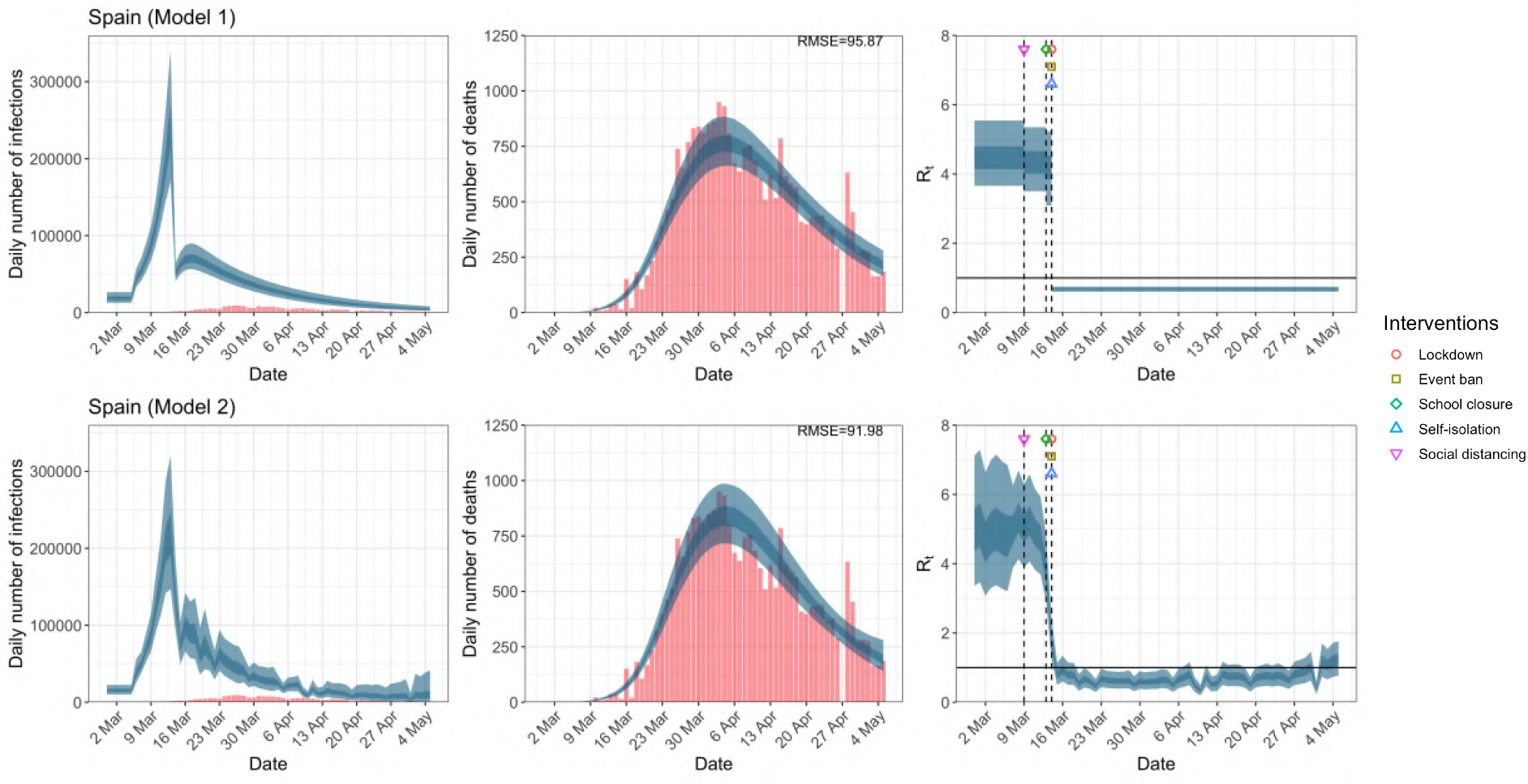
Daily infections, daily deaths and *R*_*t*_ in Spain until May 5th. The start time for the plots is 10 days before 10 deaths are recorded. Observed counts of daily infections and daily deaths are shown in red, and their corresponding 50% and 95% CIs are shown in dark blue and light blue respectively.

**Figure C.10:**
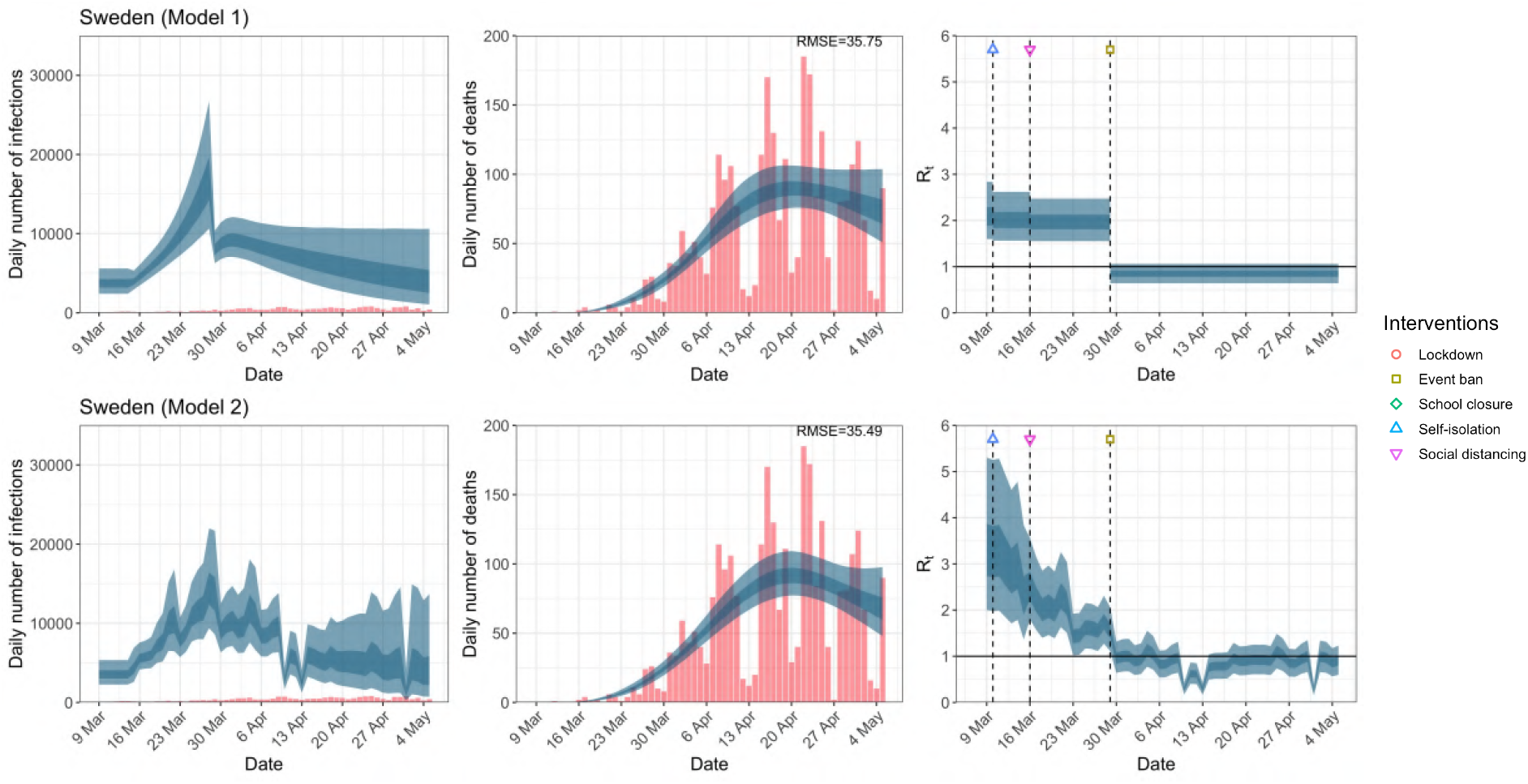
Daily infections, daily deaths and *R*_*t*_ in Sweden until May 5th. The start time for the plots is 10 days before 10 deaths are recorded. Observed counts of daily infections and daily deaths are shown in red, and their corresponding 50% and 95% CIs are shown in dark blue and light blue respectively.

**Figure C.11:**
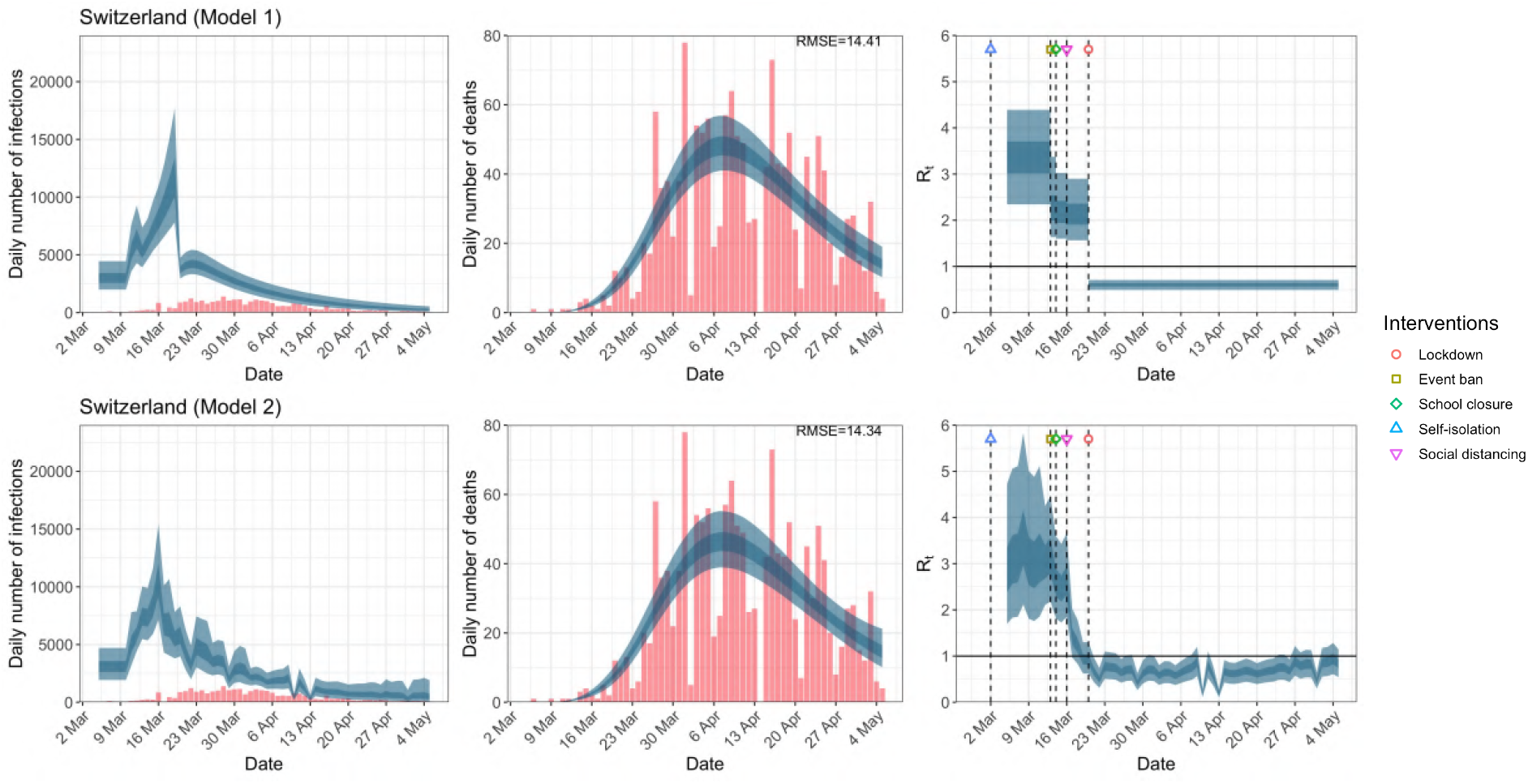
Daily infections, daily deaths and *R*_*t*_ in Switzerland until May 5th. The start time for the plots is 10 days before 10 deaths are recorded. Observed counts of daily infections and daily deaths are shown in red, and their corresponding 50% and 95% CIs are shown in dark blue and light blue respectively.

**Figure C.12:**
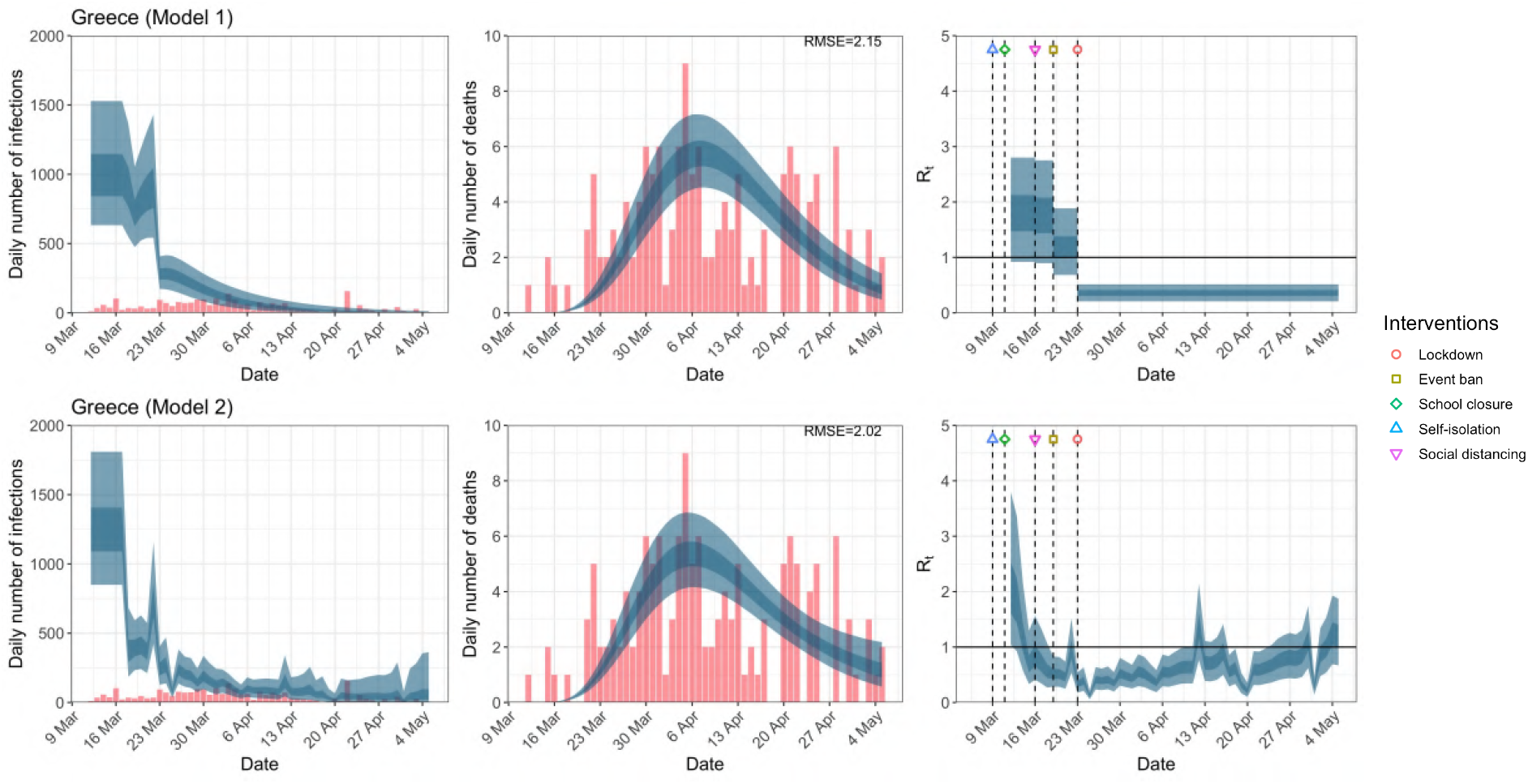
Daily infections, daily deaths and *R*_*t*_ in Greece until May 5th. The start time for the plots is 10 days before 10 deaths are recorded. Observed counts of daily infections and daily deaths are shown in red, and their corresponding 50% and 95% CIs are shown in dark blue and light blue respectively.

**Figure C.13:**
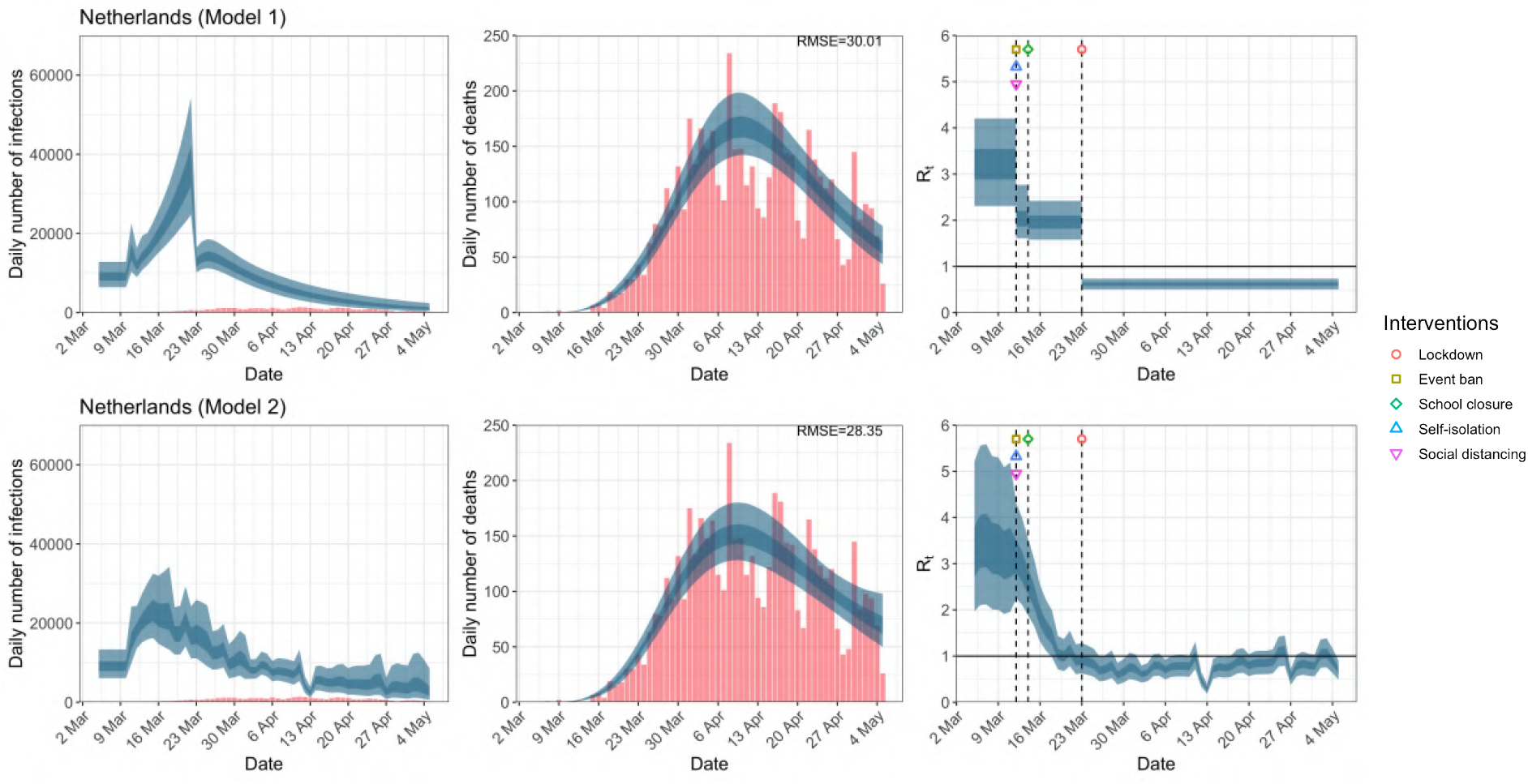
Daily infections, daily deaths and *R*_*t*_ in the Netherlands until May 5th. The start time for the plots is 10 days before 10 deaths are recorded. Observed counts of daily infections and daily deaths are shown in red, and their corresponding 50% and 95% CIs are shown in dark blue and light blue respectively.

**Figure C.14:**
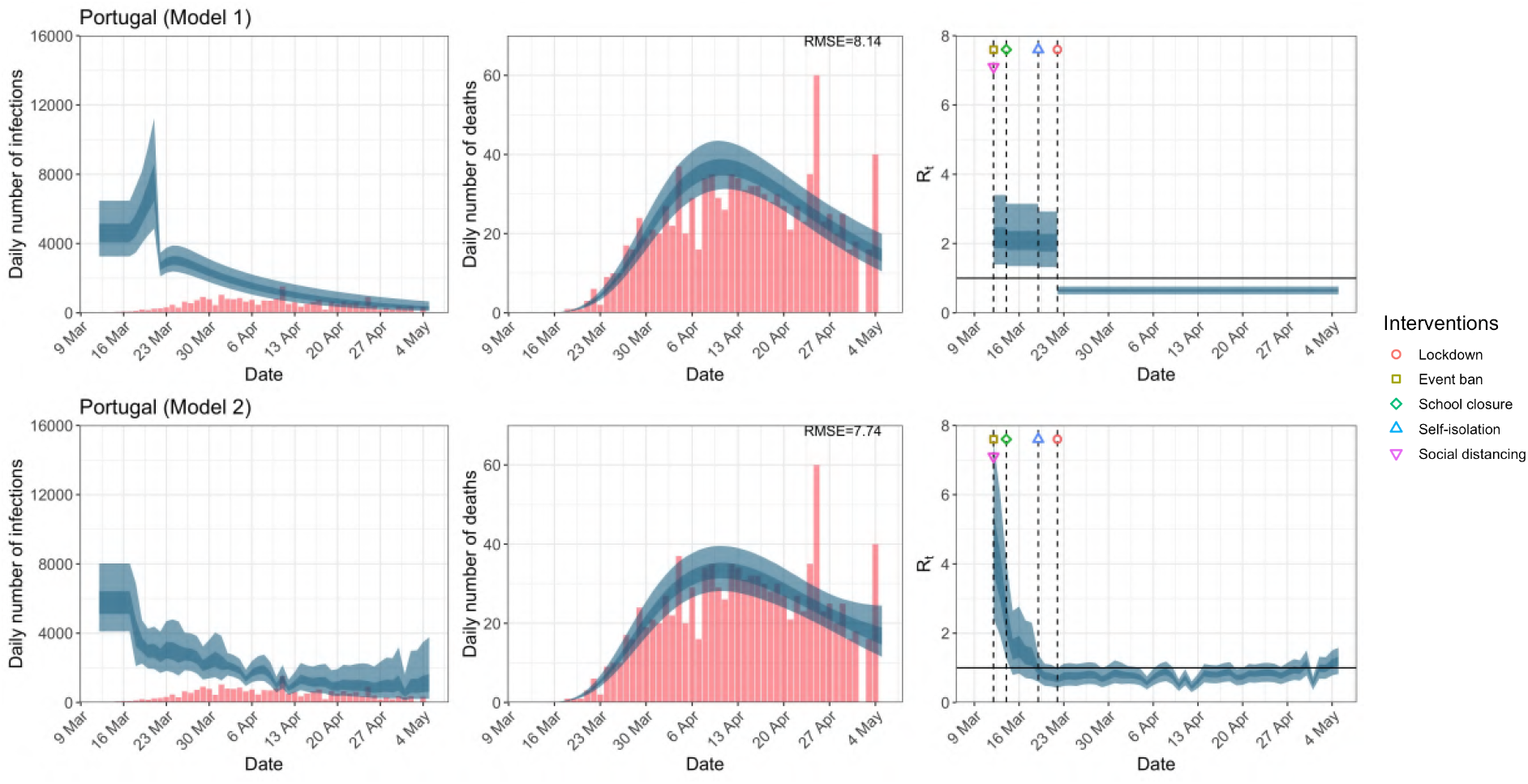
Daily infections, daily deaths and *R*_*t*_ in Portugal until May 5th. The start time for the plots is 10 days before 10 deaths are recorded. Observed counts of daily infections and daily deaths are shown in red, and their corresponding 50% and 95% CIs are shown in dark blue and light blue respectively.

**Table C.1:** Comparison of the value of *R*_*t*_ at lockdown (LD) and its 95% CIs between model 1 and 2 for all eleven countries analysed in Flaxman et al. ^1^ and an additional three countries of Greece, Netherlands and Portugal, for the time horizon March 4th to May 5th.

| Country | Model 1 |  |  | Model 2 |
| --- | --- | --- | --- | --- |
| | $R_t$ one day before LD | $R_t$ at LD | % change | $R_t$ at LD |
| UK | 3.31<br>(2.55, 3.87) | 0.68<br>(0.57, 0.80) | -79.18<br>(-84.65, -70.85) | 1.11<br>(0.74, 1.60) |
| Austria | 2.08<br>(1.17, 3.57) | 0.52<br>(0.41, 0.64) | -73.01<br>(-85.48, -57.22) | 0.87<br>(0.41, 1.55) |
| Belgium | 2.90<br>(2.06, 4.01) | 0.72<br>(0.62, 0.83) | -74.14<br>(-83.75, -61.69) | 1.46<br>(1.00, 1.99) |
| Denmark | 2.28<br>(1.39, 3.53) | 0.68<br>(0.57, 0.79) | -68.63<br>(-80.92, -51.75) | 0.57<br>(0.28, 1.04) |
| France | 3.03<br>(2.18, 4.14) | 0.75<br>(0.65, 0.84) | -74.61<br>(-83.60, -63.22) | 1.70<br>(1.16, 2.40) |
| Germany | 3.65<br>(2.90, 4.40) | 0.73<br>(0.62, 0.84) | -79.78<br>(-85.08, -72.05) | 1.02<br>(0.68, 1.44) |
| Italy | 2.11<br>(1.51, 2.86) | 0.71<br>(0.64, 0.78) | -65.31<br>(-75.81, -51.48) | 1.28<br>(0.86, 1.73) |
| Norway | 1.72<br>(0.99, 2.77) | 0.44<br>(0.28, 0.60) | -72.77<br>(-85.98, -55.88) | 0.50<br>(0.28, 0.79) |
| Spain | 4.19<br>(3.09, 5.24) | 0.68<br>(0.60, 0.75) | -83.53<br>(-87.98, -77.23) | 1.78<br>(1.22, 2.42) |
| Sweden | - | - | - | - |
| Switzerland | 2.15<br>(1.57, 2.90) | 0.60<br>(0.49, 0.71) | -71.14<br>(-82.00, -57.72) | 0.93<br>(0.62, 1.30) |
| Greece | 1.20<br>(0.68, 1.89) | 0.36<br>(0.21, 0.51) | -68.90<br>(-83.34, -48.58) | 0.34<br>(0.18, 0.54) |
| Netherlands | 1.97<br>(1.58, 2.42) | 0.62<br>(0.50, 0.73) | -68.09<br>(-78.27, -55.73) | 0.93<br>(0.63, 1.28) |
| Portugal | 2.04<br>(1.32, 2.92) | 0.65<br>(0.53, 0.76) | -66.93<br>(-79.89, -47.70) | 0.67<br>(0.42, 0.99) |

